# Outcome evaluation of the M-Mama emergency transport system (EmTS) and factors associated with its utilisation among lactating mothers in Kigoma District Council, Tanzania: A cross-sectional study

**DOI:** 10.64898/2025.12.09.25341667

**Authors:** Julius Masaba, Mackfallen G. Anasel, Laurencia Mushi

## Abstract

**Background:** Sub-Saharan Africa accounts for approximately 70% of maternal deaths worldwide. In Tanzania, the maternal mortality ratio (MMR) has decreased from 556 deaths per 100,000 live births in 2016 to 104 deaths per 100,000 live births in 2022. The major cause of these deaths is limited access to emergency obstetric care. To address this challenge, the M-Mama program was implemented to provide prompt transportation for expectant mothers experiencing obstetric complications. This evaluation study examined the factors associated with the utilization of the M-Mama Emergency Transport System (EmTS) among lactating mothers in Kigoma District Council.

**Methodology:** A cross-sectional study design was employed using a purposive sampling technique to select lactating mothers who had experienced obstetric or neonatal emergencies. Data were collected using structured questionnaires. Logistic regression analysis was performed to determine the association between independent variables and the dependent variable (utilization of the M-Mama EmTS).

**Results:** Utilization of the M-Mama EmTS was low, with only 36% of respondents reporting use of the system in Kigoma District Council. However, utilization was significantly associated with awareness of the program (AOR = 69.62; 95% CI: 25.52–189.9; p < 0.001), parity (AOR = 3.05; 95% CI: 1.15–8.12; p = 0.025), and education level (AOR = 0.36; 95% CI: 0.17–0.78; p = 0.009).

**Conclusion:** Utilization of the M-Mama Emergency Transport System was influenced by awareness of the program, parity, education level, income, occupation, and residence.

## Introduction

Maternal and newborn mortality remain pressing public health challenges globally, with the majority of deaths occurring in low- and middle-income countries [1]. Sub-Saharan Africa bears a disproportionate burden, where preventable delays in accessing skilled care continue to threaten the survival of mothers and infants [2].

The present evaluation study is linked on the Three Delays Model [4], which describes barriers to maternal health service use. The model highlights three key delays that contribute to negative outcomes: The delay in deciding to seek care (Delay I), The delay in reaching health facility (Delay II), and the delay in receiving care upon arrival (Delay III) [5]. The Three Delays Model provides a theoretical basis for examining both socioeconomic and demographic factors (age, marital status, parity, education, income, distance, occupation, residence) as determinants of M-Mama Emergency Transport System (EmTS) utilization (***Fig 1***.)

**Fig 1.**
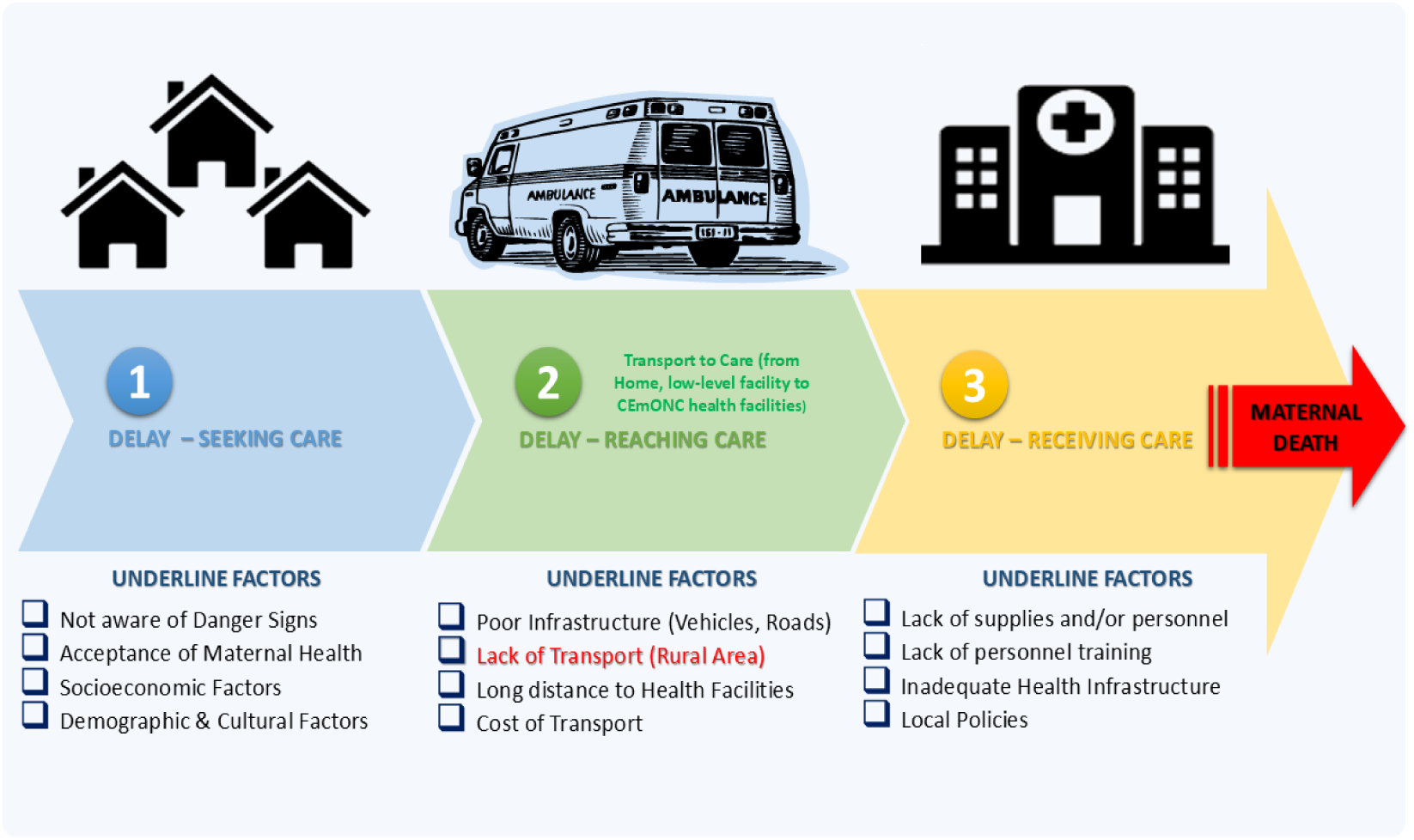
The Three Delays Model of maternal mortality showing Delay 2 (Lack of Transport)

Tanzania has experienced significantly reduction of Maternal Mortality Ratio from 556 deaths per 100,000 live births to 104 per 100,000 live births in 2022 [14, 15]. District regions such as Kigoma are particularly affected, where poor road infrastructure, long distances to health facilities, and limited ambulance coverage hinder timely access to emergency obstetric and newborn care [1]. Many women rely on bicycles, walking, or informal motorcycle during emergencies, which are often unsafe or unavailable. These transport barriers contribute to preventable maternal and neonatal deaths by reinforcing delays in reaching appropriate healthcare facilities [16, 17].

To address these challenges, the Government of Tanzania, in collaboration with partners including Vodafone Foundation, USAID, and Touch Foundation, Pathfinder International, implemented the M-Mama Emergency Transport System (EmTS) in Shinyanga region for pilot and now to all region of Tanzania [11]. The program integrates a toll-free emergency hotline with a digital dispatch system that connects women in need with either ambulances or trained community drivers when ambulances are not available [8]. Community drivers are reimbursed through mobile money to ensure reliability, while women are transported to the nearest health facility capable of managing obstetric emergencies. By addressing the second delay reaching appropriate care the M-Mama Emergency Transport System (EmTS) represents a community-based innovation designed to reduce maternal and newborn deaths in underserved areas [11]. The M-Mama program has been shown to reduce Delay II and improve timely referral transport, thereby increasing facility-based births and contributing to reductions in maternal and neonatal mortality [8]. Similarly, [9] found that the M-Mama Champion program enhanced awareness of obstetric danger signs, birth preparedness, and complication readiness by over 60% among pregnant women in Dodoma.

The conceptual model of this study was developed by mapping the independent variables socioeconomic and demographic factors (age, marital status, parity, education, income, distance, occupation and residence) and project enabling factors (M-Mama awareness, M-Mama Advert, M-Mama Source of Advert, M-Mama number reachable and emergency obstetric and neonatal care availability) onto Delay I and Delay II of the Three Delays Model (***Fig 2***.)

**Fig 2.**
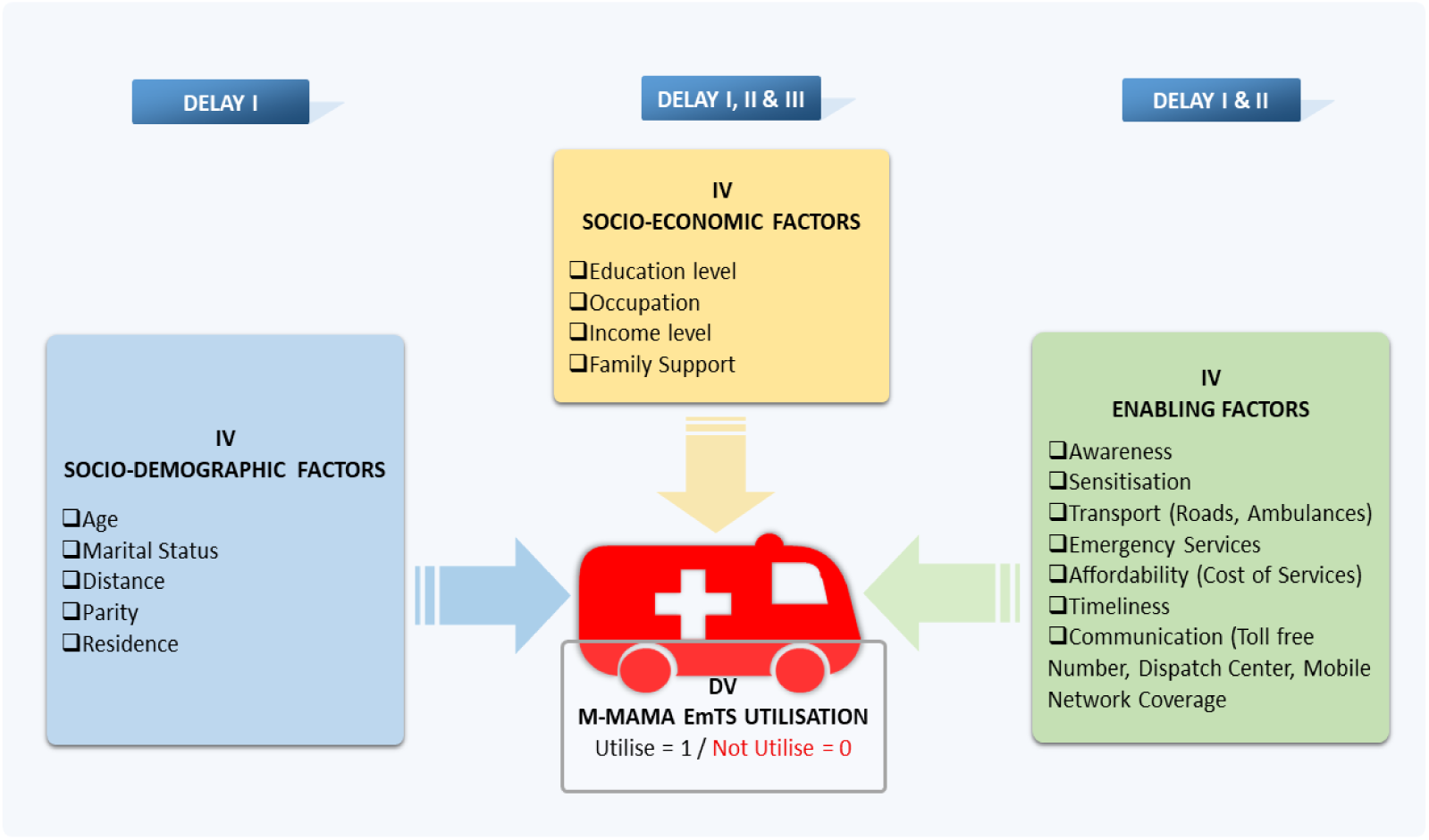
Conceptual framework for factors associated with utilization of the M-Mama Emergency Transport System (EmTS).

Despite evidence that the M-Mama Emergency Transport System (EmTS) reduces transportation barriers and improves referrals, existing studies have focused largely, lessons learnt for best practices, implementation and cost-effectiveness in pilot regions such as Shinyanga [9, 11, 12]. Less is known about the determinants of mothers’ actual utilization of M-Mama, particularly how socioeconomic, demographic, and awareness-related factors (Delay I) interact with project-enabling mechanisms to influence the utilization of M-Mama Emergency Transport System (EmTS). Moreover, evidence from Kigoma District Council remains scarce, where sociocultural and geographical dynamics may shape utilization differently. The current evaluation study therefore seeks to addresses this gap by evaluating the M-Mama Emergency Transport System (EmTS) and factors associated with its utilization in Kigoma District Council, Tanzania.

## MATERIALS AND METHODS

### Study Area and Period

The Kigoma District Council in Tanzania’s Kigoma Region was the site of the evaluation study. Covering an area of 967.7 square Kilometers, Kigoma District Council is located in north-western Tanzania along the eastern shore of Lake Tanganyika, with a population of 222,792, including 104,903 males and 117,889 females at the time of the study [18] ***(Fig 3*.)**. The evaluation study was conducted from November 2024 to September 2025 covering four seasons: Proposal development (November 2024 to February 2025), Pilot, training research assistant and any preparation activities (March 2025 to July 2025), Recruitment period and data collection were conducted from (23/07/2025 to 14/08/2025), Analysis, Interpretation, and Presentation of results (August to September 2025).

**Fig 3.**
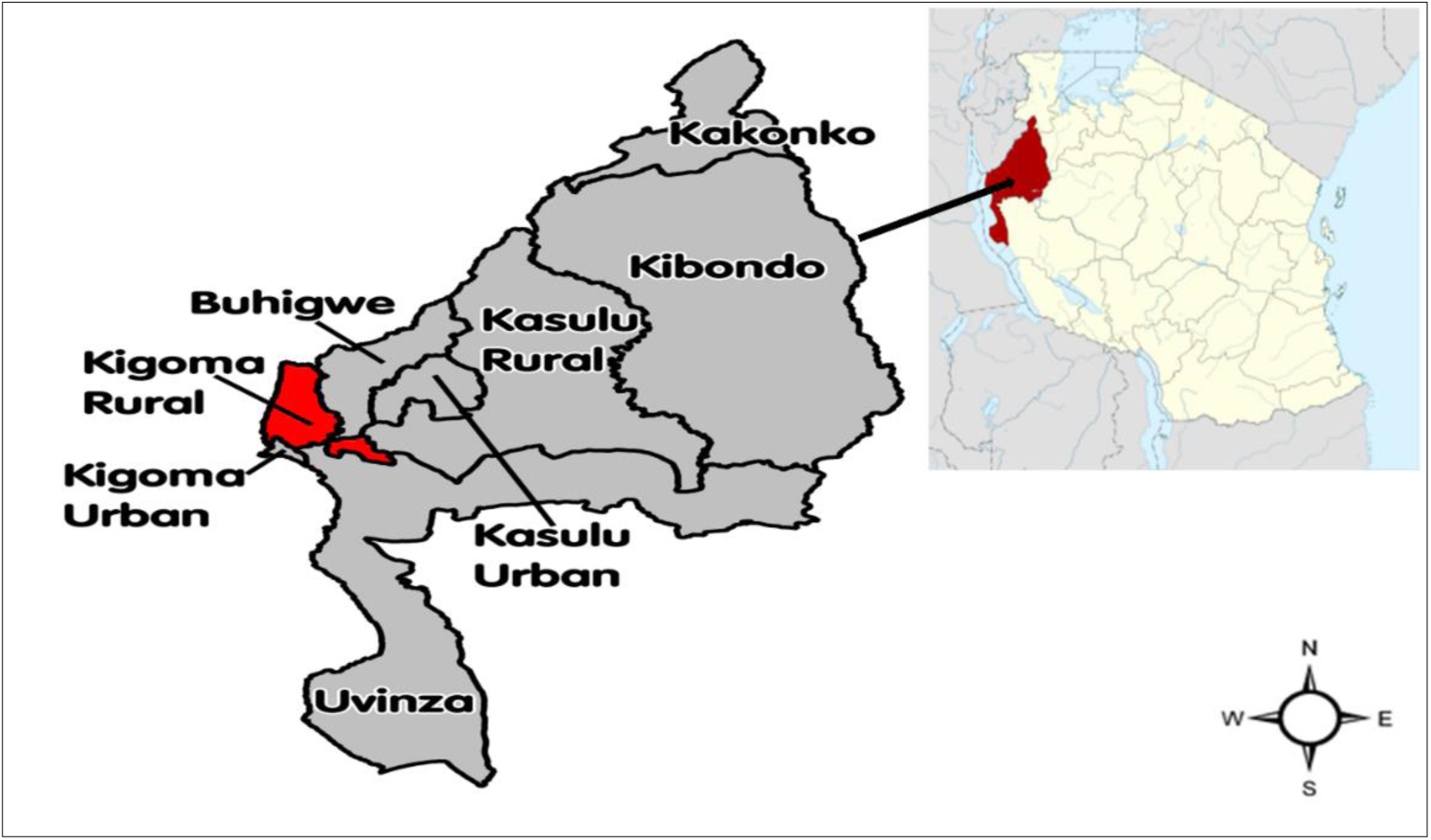
Map of Kigoma District Council. A Geographic representation of the study location within Kigoma District Council.

### Study Approach and Design

The evaluation study used a quantitative and descriptive facility-based cross-sectional study design to examine factors influencing the utilization of the M-Mama Emergency Transport System (EmTS) in Kigoma District Council. This design allows for the collection of precise data at a single point in time, providing a snapshot of utilization patterns and associated factors [19].

### Study Population and Criteria

The evaluation study population consisted of lactating mothers with children under two years old, who had experienced an obstetric or neonatal emergency during their pregnancy.

The inclusion criteria comprised lactating mothers of reproductive age (15–49 years) with children under two years, specifically those who gave birth between January 2023 to December 2024, experienced an obstetric or neonatal emergency, residents of Kigoma District Council for not less than six months and were attending a facility’s Reproductive and Child Health (RCH) services. In selecting health facilities, priority was given to those located in hard-to-reach areas with poor transportation infrastructure, as these are the primary users of the M-Mama EmTS; a total of 21 facilities were purposively selected out of 50 health facilities in Kigoma District Council.

The exclusion criteria included mothers who did not meet the age range or child criteria, had not given birth in 2023 or 2024, had not experienced an obstetric or neonatal emergency, or were not residents of Kigoma District Council for less than six months. Additionally, mothers who were unable to provide informed consent or had severe health conditions were excluded.

### Variables

The dependent variable was the utilization of M-Mama (EmTS) while the Independent variables including Socioeconomic and Demographic factors such as (age, parity, marital status, education, income, distance, residence and occupation), project-enabling factors (M-Mama awareness, M-Mama advertisements, M-Mama sources of advertisements, accessibility of the M-Mama number, and availability of emergency obstetric and neonatal care) and Perception (Satisfactions, Effectiveness, Trust, Quick response and Recommendation); When combined, these factors allow for a thorough examination of M-Mama (EmTS) service utilization.

### Sample Size

The sample size for this quantitative study was determined by using Yamane’s Formula as follows: Yamane’s Formula n = N/(1+N*e2) Whereby n = Required sample size, N = 1,473 Lactating mothers who experienced obstetric or neonatal emergencies having children; Therefore n = 315.

### Sampling Procedure

This evaluation study used a purposive sampling technique to select 21 health facilities in hard-to-reach areas targeted by the M-Mama Emergency Transport System (EmTS) programme, as semi-urban facilities had better access to transport and emergency care. From each selected 21 facility, 15 lactating mothers with the required criteria were purposively recruited.

### Source of Data

The data for this evaluation study were drawn from two sources: primary data obtained through structured questionnaires administered to lactating mothers using the Kobo tool, and secondary data from documentary reviews, including M-Mama EmTS program reports, DHIS2, and related studies on Maternal health utilization, obstetric and neonatal emergencies. Using both sources strengthen validity and reliable data [20].

### Data Collection Methods and Tools

A structured questionnaire with closed-ended questions was designed to capture demographic, socioeconomic, Perception and project-enabling factors influencing the utilization of the M-Mama Emergency Transport System (EmTS), ensuring consistency and ease of data analysis [20], [21]. Data were collected electronically using the Kobo Collect application, which enables real-time entry, minimizes errors associated with manual data entry, and improves data quality. Before administered face-to-face with lactating mothers attending RCH health facilities who met the inclusion criteria, providing comprehensive information on M-Mama EmTS utilization the questionnaire was prepared in English and translated to the local language (Swahili) and back-translated to English by language experts to see the consistency and validity.

### Data Analysis

The evaluation study applied quantitative data analysis using STATA 17. The dataset was cleaned to address issues such as missing values and outliers. Descriptive statistics, including frequencies and percentages, were used to summarized key variables. Cross-tabulations were used to explore associations between categorical variables, ensuring the minimum expected count requirement was met. For inferential analysis, the chi-square test was employed to examine the relationship between the binary dependent variable utilization of the M-Mama Emergency Transport System (EmTS) and categorical independent variables [22]. Since both dependent and independent variables were categorical, the chi-square test assessed whether observed proportional differences were statistically significant [23]. To identify determinants of M-Mama Emergency Transport System (EmTS) utilization, binary logistic regression model was used, as the dependent variable was binary (1 = utilise, 0 = not utilise) [24]. Logistic regression assumptions including linearity of predictors with the logit, independent observations, absence of multicollinearity, and sufficient sample size were considered [25]. Odds ratios (ORs) were calculated to estimate the strength of associations between predictors and utilization. Model fit and logit linearity were assessed using the Hosmer-Lemeshow goodness-of-fit test. Below is the Model description of the logit function [26].

Logit 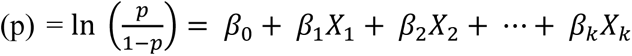; Whereby p = The probability that a Lactating Mother utilizes M-Mama EmTS, *β*_0_ = Is the intercept, *β*_1_, *β*_2_………, *β*_*k*_= Regression Coefficients, *X*_1_, *X*_2_………, *X*_*k*_ = Independent Variables.

The coefficients CI were exponentiated to obtain odds ratios (ORs), which indicate the change in odds of utilizing the M-Mama Emergency Transport System (EmTS) associated with a one-unit change in each independent variable, holding other variables constant. This approach provided meaningful insights into factors influencing M-Mama (EmTS) utilization in Kigoma District Council.

### Ethical Consideration

Ethical approval to conduct this evaluation study was obtained from:

1. Office of Deputy Vice Chancellor (Academic, Research and Consultancy), Mzumbe University, Morogoro Region, Tanzania *– Approval granted, Letter Ref. No. MU/DPGS/INT/38/Vol.IV/592 of March 14, 2025*.
2. Office of District Executive Director (DED), Kigoma District Council, Tanzania *– Approval granted, Letter Ref. No. KDC/H6/1/1/1 of May 13, 2025*.
3. Office of District Executive Director (DED), Kigoma District Council, Tanzania to health Facilities (Introduction and Permission to conduct data collection to 21 health facilities of Kigoma District Council) – *Approval granted, Letter Ref. No. KDC/H1/10 of May 13, 2025*.
4. Written informed consent was provided to participants before data collection, either by signing or fingerprinting depending on literacy, after being briefed in the local language on the study’s purpose, procedures, risks, and benefits [29]. Consent forms were stored separately from the study data, which contained no personal identifiers and was fully anonymised for analysis.

## RESULTS

### Demographic and Socioeconomic Characteristics of Respondents

Among the 315 participants, 58% were aged 15–25 years, 62% had 1–2 children, and 83% were married. A majority (68%) reported low household income, while 53% had no formal education. Regarding occupation, 55% were farmers, and 61% resided in villages. Furthermore, 68% of participants lived 4–6 km from the nearest RCH facility (Table 1).

**Table 1.** Socioeconomic and demographic characteristics of respondents, Kigoma District Council, Tanzania, 2025 (n = 315).

| Variables | Frequency (n) | Percentage (%) |
| --- | --- | --- |
| **Age** |  |  |
| 15-25 | 184 | 58 |
| 26-36 | 131 | 42 |
| **Parity** |  |  |
| 1-2 | 194 | 62 |
| 3-4 | 121 | 38 |
| **Marital Status** |  |  |
| Single | 52 | 17 |
| Married | 263 | 83 |
| **Income Level** |  |  |
| Low Income | 214 | 68 |
| High Income | 101 | 32 |
| **Education Level** |  |  |
| None | 166 | 53 |
| Primary | 109 | 34 |
| Secondary+ | 40 | 13 |
| **Occupation** |  |  |
| Farmer | 172 | 55 |
| Tailor | 68 | 21 |
| Housewife | 44 | 14 |
| Employed | 31 | 10 |
| **Residence** |  |  |
| Village | 191 | 61 |
| Town | 124 | 39 |
| **Distance to RCH** |  |  |
| 0-3 Km | 117 | 37 |
| 4-6 Km | 198 | 63 |

### Socioeconomic and demographic factors associated with Utilisation of the M-Mama Emergency Transport System (EmTS)

#### Bivariable Logistic Regression Results

A bivariable logistic regression model was fitted to examine the crude associations between each independent variable and the utilization of the M-Mama Emergency Transport System (EmTS). The results are shown as crude odds ratios (CORs) with 95% confidence intervals (CIs). Variables with a *p*-value of ≤ 0.25 were included in the multivariable model (Table 2). As illustrated in Table 2, mothers with 3–4 children were significantly less likely to utilise the M-Mama EmTS compared to those with 1–2 children (COR = 0.22; 95% CI: 0.13–0.38; *p* = 0.001).

**Table 2.** Bivariable and Multivariable analysis showing associations between independent variables and utilization of M-Mama EmTS in Kigoma District Council, Tanzania, 2025 (n = 315).

| Variables | M-Mama Utilisation |  | Crude OR<br>(95% CI) | P - Value | Adjusted OR<br>(95% CI) | P - Value |
| --- | --- | --- | --- | --- | --- | --- |
|  | Utilise | Not utilise |  |  |  |  |
| Age |  |  |  |  |  |  |
| 15-25 | 66 (59) | 118 (58) | 1.00 |  |  |  |
| 26-36 | 46 (41) | 85 (42) | 0.97 (0.61, 1.55) | 0.890 |  |  |
| Parity |  |  |  |  |  |  |
| 1-2 | 92 (82) | 102 (50) | 1.00 |  |  |  |
| 3-4 | 20 (18) | 101 (50) | 0.22 (0.13, 0.38) | 0.001 | 0.32 (0.17, 0.60) | 0.001 |
| Marital Status |  |  |  |  |  |  |
| Married | 18 (16) | 34 (17) | 1.00 |  |  |  |
| Single | 94 (84) | 169 (83) | 0.95 (0.51, 1.78) | 0.653 |  |  |
| Income Level |  |  |  |  |  |  |
| Low Income | 67 (60) | 147 (72) | 1.00 |  |  |  |
| High Income | 45 (40) | 56 (28) | 1.76 (1.08, 2.87) | 0.023 | 2.71 (1.34, 5.47) | 0.005 |
| Education Level |  |  |  |  |  |  |
| None | 78 (70) | 88 (43) | 1.00 |  |  |  |
| Primary | 25 (22) | 84 (41) | 0.34 (0.20, 0.58) | 0.001 | 0.16 (0.08, 0.33) | 0.001 |
| Secondary+ | 9 (8) | 31 (15) | 0.33 (0.15, 0.73) | 0.006 | 0.12 (0.39, 0.37) | 0.001 |
| Occupation |  |  |  |  |  |  |
| Farmer | 58 (52) | 114 (56) | 1.00 |  |  |  |
| Tailor | 28 (25) | 40 (20) | 1.38 (0.77, 2.45) | 0.279 |  |  |
| Housewife | 18 (16) | 26 (13) | 1.36 (0.69, 2.68) | 0.374 |  |  |
| Employed | 8 (7) | 23 (11) | 0.68 (0.29, 1.62) | 0.389 |  |  |
| Residence |  |  |  |  |  |  |
| Village | 82 (73) | 109 (54) | 1.00 |  |  |  |
| Town | 30 (27) | 94 (46) | 0.42 (0.26, 0.70) | 0.001 | 0.51 (0.25, 1.04) | 0.064 |
| Distance to Reproductive Child Health |  |  |  |  |  |  |
| 4-6 Km | 26 (23) | 91 (45) | 1.00 |  |  |  |
| 0-3 Km | 86 (77) | 112 (55) | 0.37 (0.22, 0.63) | 0.001 | 0.37 (0.18, 0.57) | 0.007 |

Mothers with higher income were 1.76 times more likely to utilize the M-Mama EmTS compared to those with low income (COR = 1.76; 95% CI: 1.08–2.87; *p* = 0.023) (Table 2).

In terms of education, lactating mothers with primary education (COR = 0.36; 95% CI: 0.19–0.58; *p* = 0.001) and those with secondary education or higher (COR = 0.33; 95% CI: 0.15–0.73; *p* = 0.006) were significantly less likely to use the service compared to those with no education. Similarly, mothers living in towns were less likely to utilise the M-Mama Emergency Transport System (EmTS) than those living in villages (COR = 0.42; 95% CI: 0.26–0.70; *p* = 0.001) (Table 2). Notably, awareness was the strongest predictor mothers aware of the M-Mama EmTS were 36 times more likely to utilise it than those who were not aware (COR = 36.2; 95% CI: 18.7–70.2; *p* = 0.001) (Table 2).

#### Multivariable Logistic Regression Results

After adjusting for potential confounders, several variables remained significant predictors of M-Mama EmTS utilization. High parity continued to show a strong influence on utilization (AOR = 0.73; 95% CI: 0.42–1.28; *p* = 0.001). Similarly, mothers with higher income were 2.71 times more likely to use the M-Mama services compared to those with lower income (AOR = 2.71; 95% CI: 1.34–5.47; *p* = 0.005) (Table 2).

Educational level was also found to reduce the likelihood of service utilization. Mothers with primary education had significantly lower odds of using the service compared to those with no education (AOR = 0.16; 95% CI: 0.08–0.33; *p* = 0.001). Likewise, those with secondary education and above were even less likely to use the service (AOR = 0.12; 95% CI: 0.04–0.37; *p* = 0.001) (Table 2).

Distance to facility was also significant: mothers living within 0–3 km was less likely to utilize the service compared to those living within 4–6 km (AOR = 0.37; 95% CI: 0.18–0.76; *p* = 0.007) (Table 2). Residence showed an inverse relationship, with mothers living in towns being less likely to use the M-Mama EmTS compared to those living in villages (AOR = 0.51; 95% CI: 0.25–1.04; *p* = 0.064). Regarding distance, mothers living within 0–3 km of the nearest facility were significantly less likely to utilize the service compared to those living 4–6 km away (AOR = 0.37; 95% CI: 0.18–0.76; *p* = 0.007) (Table 2).

#### Proportion of Utilization of the M-Mama EmTS among Lactating Mothers

This objective aimed to describe the proportion of M-Mama Emergency Transport System (EmTS) utilisation and its characteristics among lactating mothers in Kigoma District Council.

Out of the 315 surveyed mothers, 112 (36%) reported utilising the free M-Mama EmTS during maternal emergencies, while 203 (64%) did not use the service and instead relied on other paid means of transportation illustrates the distribution of utilization of the M-Mama EmTS among lactating mothers in Kigoma District Council.

**Fig 4.**
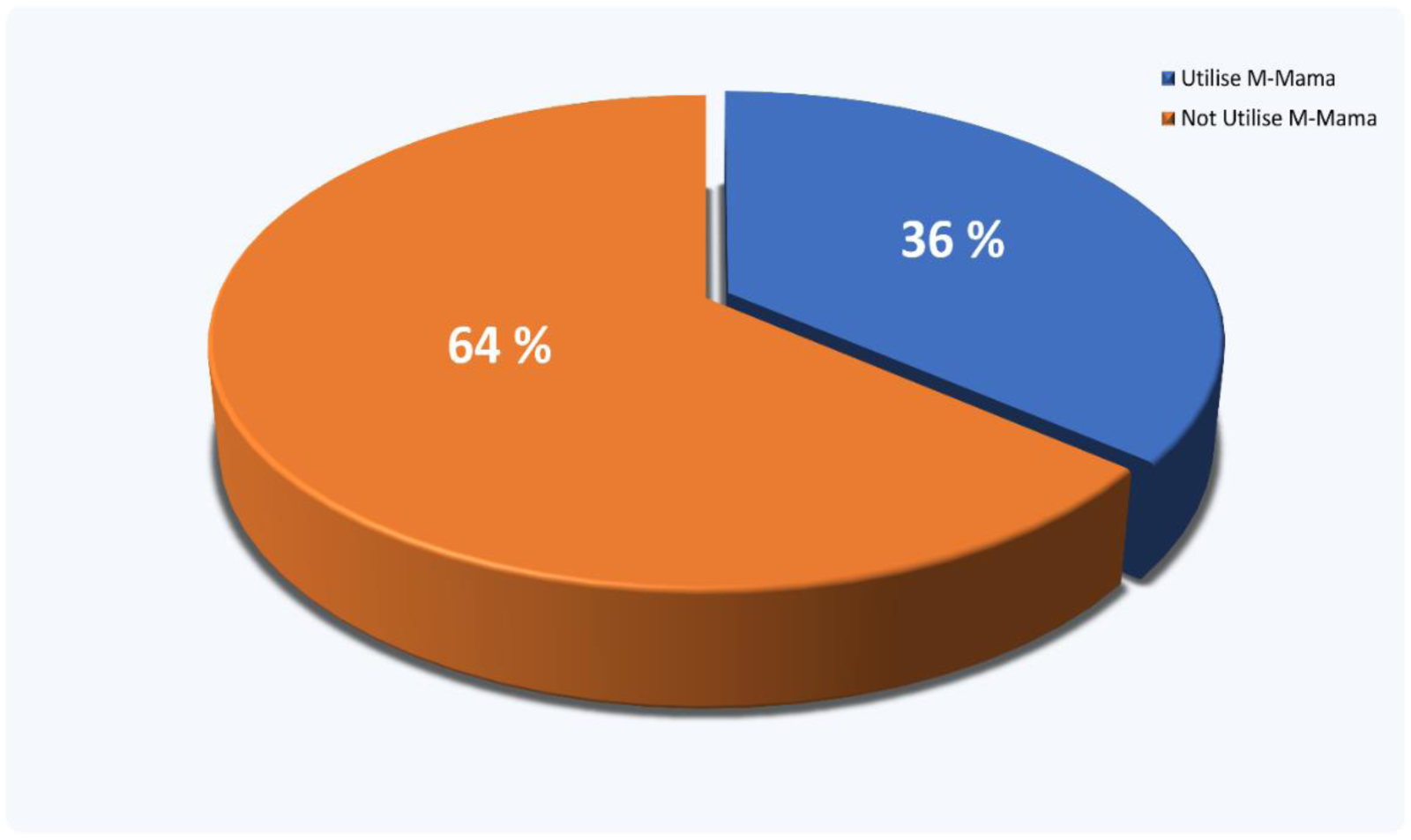
Proportion of M-Mama (EmTS) utilization among lactating mothers in Kigoma District Council, Kigoma Region, Tanzania, 2025.

Moreover, the data indicated that during obstetric and neonatal emergencies, the majority of lactating mothers 203 (65%) relied on private transport. In contrast, 77 (24%) used the M-Mama Ambulance, and 35 (11%) used the M-Mama Community Taxi. Consequently, a total of 112 lactating mothers utilized the M-Mama EmTS between 2023 and 2024, highlighting the contribution of the program in improving access to timely emergency transport ***(Fig 5.)***

**Fig 5.**
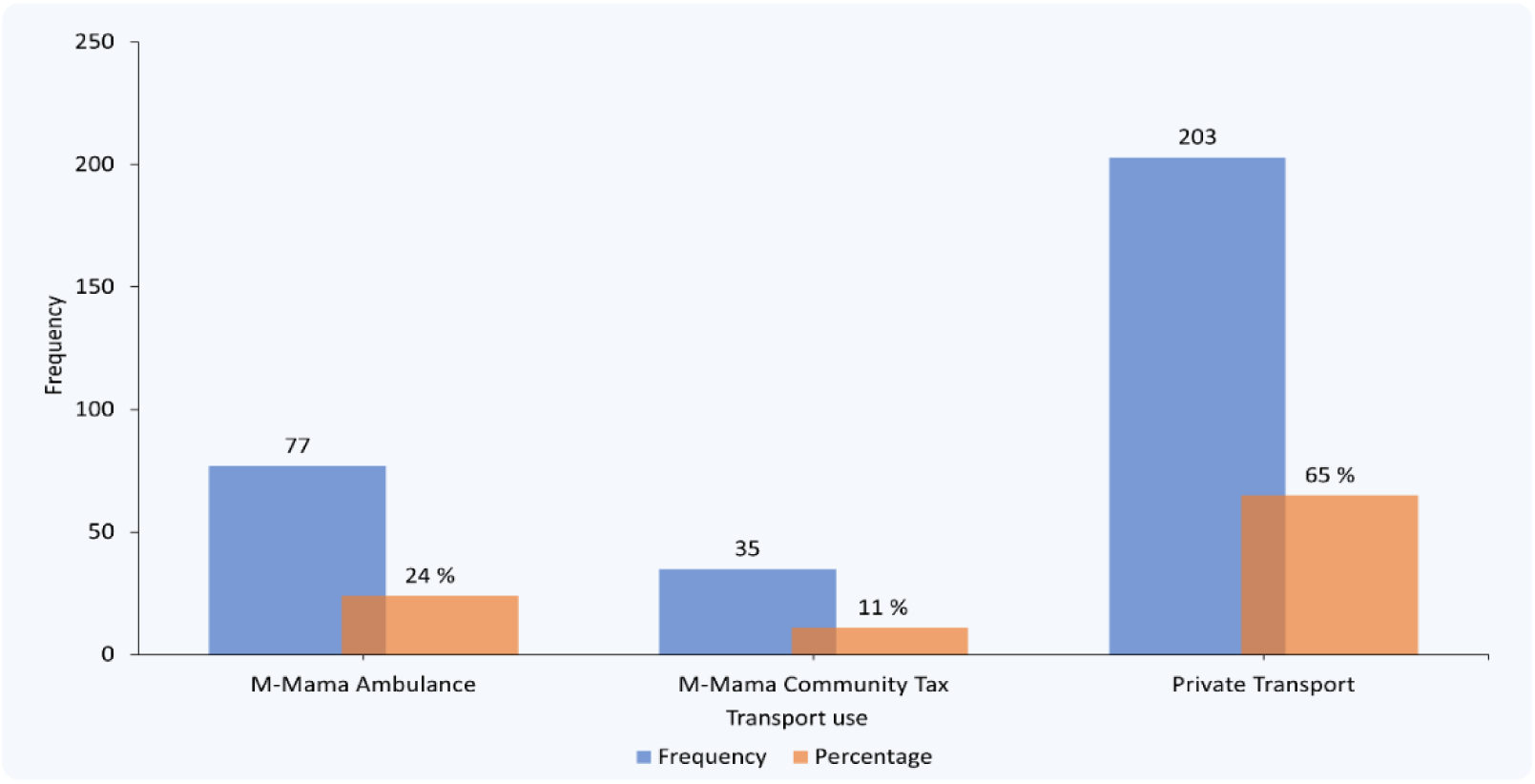
Illustrating the different transport options chosen by respondents.

Additionally, findings revealed that 42% of lactating mothers reported that the emergency circumstances prevented them from using the M-Mama EmTS, while 22% indicated that they were not aware of the service. Conversely, 20% of mothers reported utilizing the M-Mama EmTS because it is a free service, and 16% cited its quick response time as the reason for utilizing ***(Fig 6)***

**Fig 6.**
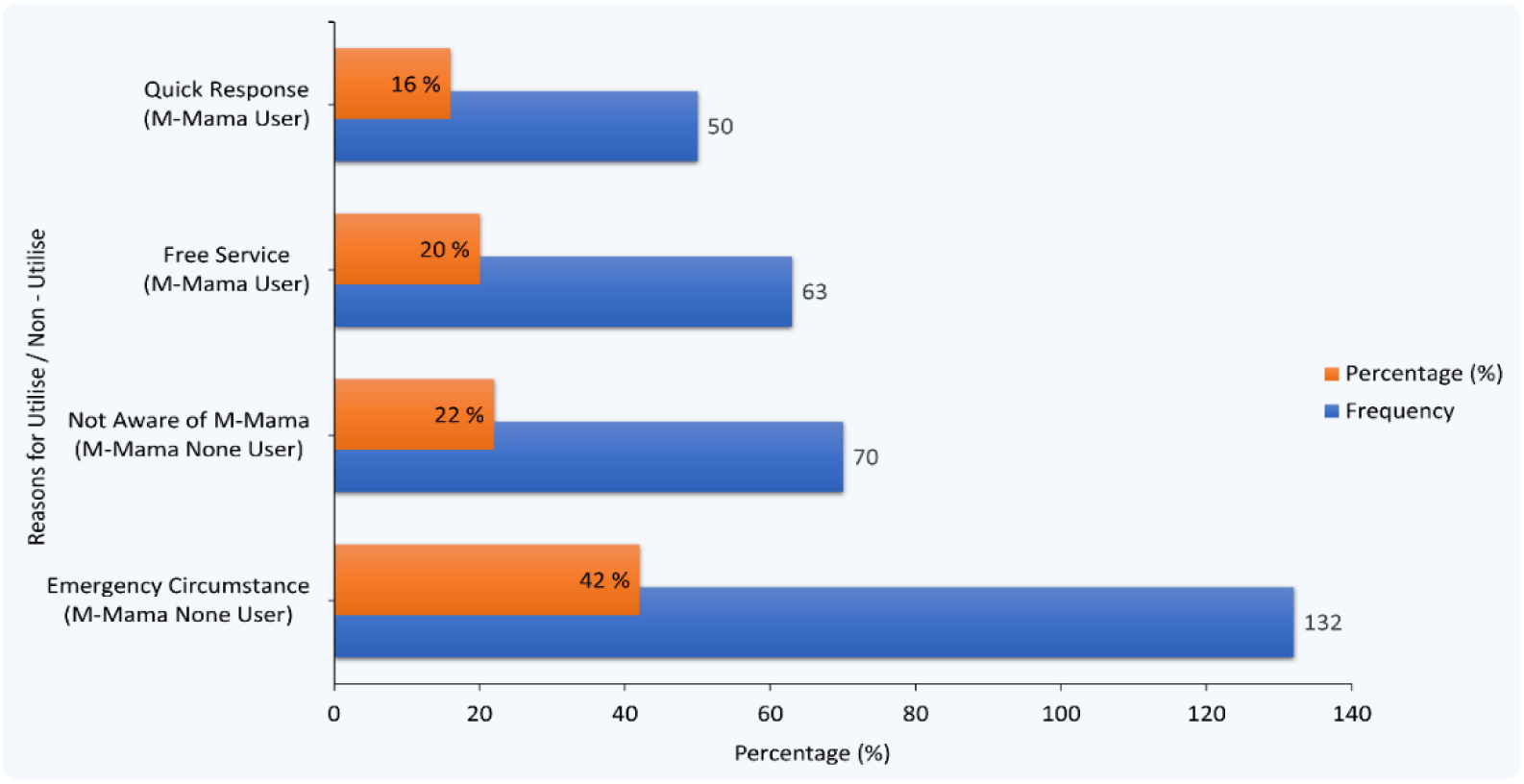
Reasons for utilise and non-utilise M-Mama Emergency Transport System (EmTS)

A cross-tabulation was conducted to assess the utilization of the M-Mama EmTS across different health facilities in Kigoma District Council. Utilization was highest at Chankele (93.3%) and Bubango (6.7%) among mothers surveyed at these facilities. In contrast, no utilization was reported at Kiganza, Kimbwela, Mwamgongo, and Pamila facilities during the survey period (0.0%) ***(Fig 7)*.**

**Fig 7.**
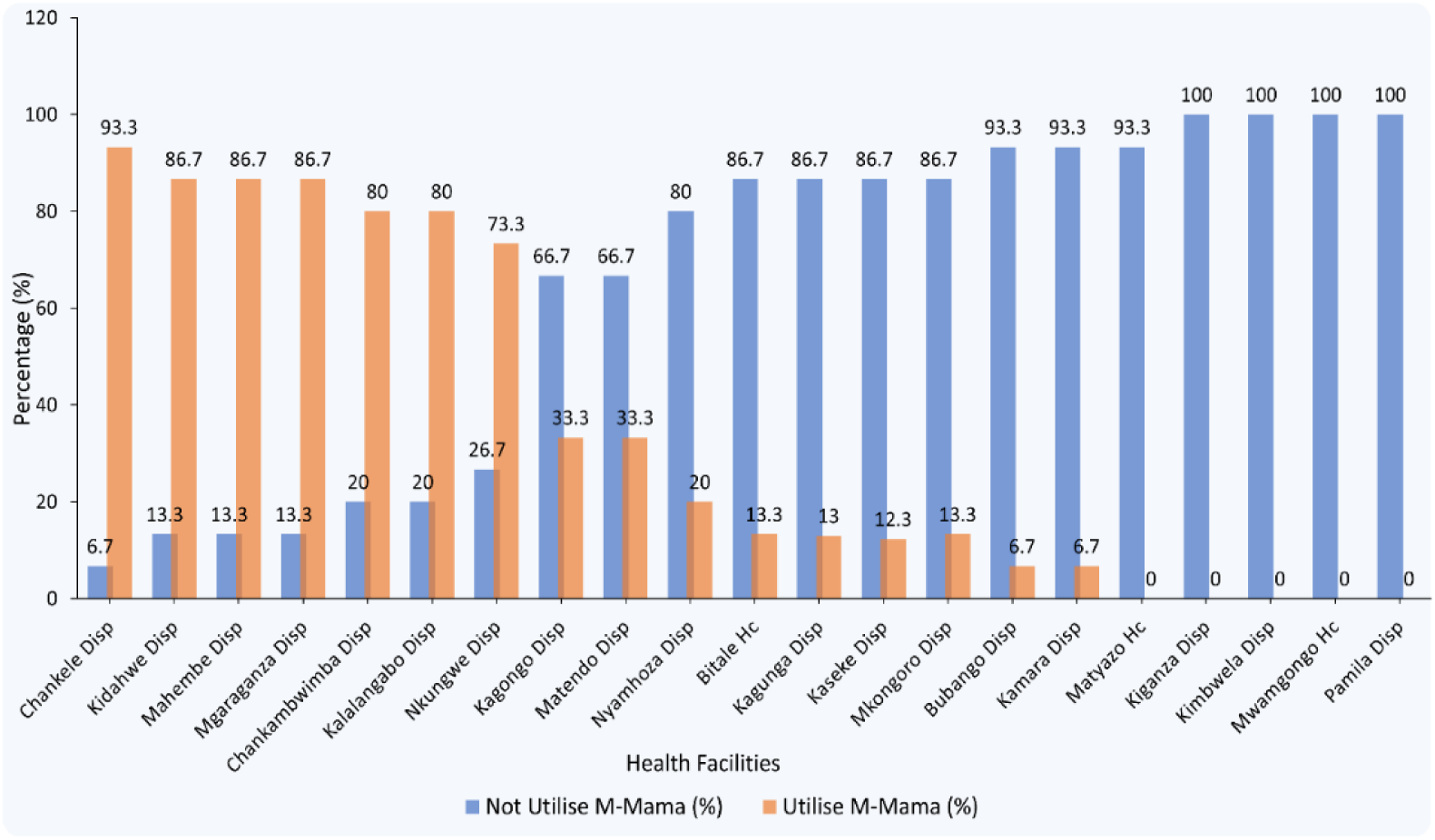
Proportion of M-Mama (EmTS) utilization across health facilities in Kigoma District Council, Kigoma Region, Tanzania, 2025.

## DISCUSSION

### Socioeconomic and demographic factors associated with Utilization of M-Mama Emergency Transport System (EmTS)

The study revealed that mothers with higher parity were less likely to utilize M-Mama EmTS compared to primiparous mothers (Table 2). This is consistent with findings from Chelia District, Oromia, where multigravida mothers were 4.8 times more likely to use ambulance services compared to primigravida women [32]. Higher parity may increase women’s awareness of obstetric complications, improve their confidence in decision-making, and strengthen their recognition of the importance of timely emergency transport.

Mothers with higher income were more likely to utilize the M-Mama EmTS compared to those with low income (Table 2). Higher-income mothers may have better access to information about M-Mama services through media, mobile phones, social networks, or health education thus are more likely to utilise M-Mama EmTS compared to low-income mothers; However, this variable was not identified as significant predictors of M-Mama EmTS utilisation. Unlike the findings in Buno Bedele Zone, Southwest Ethiopia, where low-income families were more dependent on free ambulance services and wealthier households preferred private transport [33]. These results underscore the importance of financial accessibility in overcoming the Delay II related to transportation costs.

In terms of education, lactating mothers with primary education and those with secondary education or above were significantly less likely to utilize the M-Mama EmTS compared to mothers with no education (Table 2). This may be because educated mothers often prefer private transport options, such as taxis, motorcycles, or personal cars, which they perceive as faster and more reliable. Additionally, higher education is frequently associated with formal employment, which often involves living closer to towns with better access to alternative transportation, whereas mothers with no education are more likely to reside in villages and depend on services like M-Mama for emergency transport [34]. Unlike the findings were reported in Southwest Ethiopia, where mothers who had formal education were more likely toward utilizing ambulance services compared with mothers with no formal education [33]. These results suggest that health education and awareness campaigns should target all education levels, emphasizing the safety, reliability, and efficiency of M-Mama EmTS.

Mothers residing closer to health facilities had less likelihood of using M-Mama EmTS, compared to those living within 4–6 km (remote or geographically difficult areas) (Table 2). Geographic access modeling in Kigoma Region reported that nearly one-third of births occur in locations needing a motorized transport, while another one-third still face challenge of travel times exceeding 2 hours regardless of transport mode, underscoring substantial reliance on emergency transport system like M-Mama [16]. However, deaths that are direct causes of maternal mortality were strongly associated by distance [34] This finding also reflects the “second delay” in accessing transportation and aligns with studies in District Ethiopia, which showed that poor road networks and long distances were significant barriers to ambulance utilization [27, 32, 35]. In Kigoma District Council, mountainous terrain and dispersed settlements may further exacerbate these challenges [16].

Residence exhibited an inverse relationship with utilization of the M-Mama EmTS: mothers living in towns were less likely to use the service compared to those in villages. Similarly, distance played a significant role mother residing within 0–3 km of the nearest facility was significantly less likely to use the service compared to those living 4–6 km away (Table 2). This suggests that mothers living closer to facilities or in semi urban settings may rely on walking or private modes of transport, whereas those farther away or in District areas depend more on organized emergency transport [34]. Comparable findings were reported in a study from Jimma City, Ethiopia, where patients residing within 15 km of the hospital had significantly lower odds of ambulance utilization underscoring how proximity diminishes the reliance on formal transport systems [36].

### Proportion of Lactating Mothers in the Utilization of M-Mama Emergency Transport System (EmTS)

The survey findings from Kigoma District Council indicate that a relatively small proportion of lactating mothers utilize the M-Mama EmTS, despite its free availability ***(Fig 4)***. Such limited utilisation may be attributed to several factors, including inadequate community awareness of the service, challenges in transport system readiness, and infrastructural limitations that hinder rapid access to emergency transport in District settings, [37, 38, 40–42]. Similar patterns have been documented in comparable settings: for instance, in six councils of the Shinyanga Region, 69.9% of emergency referrals utilized the EmTS, compared to just 30.1% that relied on conventional ambulance services highlighting a greater utilisation in District areas where traditional ambulances were less accessible [11]. Study in Mt. Elgon Sub-County in Kenya, where despite general awareness of free maternal healthcare policies like the Linda Mama program, uptake remained below expectations owing to gaps in infrastructure, health worker capacity, and community-level sensitization [40, 42]. Barriers have been reported in other low-resource contexts, where community-based ambulance schemes often face underutilization due to low awareness, cultural preferences, and competing private transport options [33]. Furthermore, studies from Ethiopia and Nigeria highlight that the maturity of emergency referral systems, community trust, and integration with local health structures significantly influence the adoption of free ambulance services [33, 36]. Involving communities in health education is another way to raise awareness of the services that are available and promote their utilisation. According to the World Health Organization access to healthcare services does not ensure their use; rather, system responsiveness, trust, and knowledge are crucial [43].

The survey data from Kigoma District Council show that during obstetric and neonatal emergencies, the majority of lactating mothers 65% relied on private transport, while 24% used the M-Mama Ambulance and 11% used the M-Mama Community Taxi. Altogether, 112 lactating mothers utilized the M-Mama EmTS between 2023 and 2024, underscoring the program’s role in expanding access to timely emergency transport in this setting, (Evaluation Field Data, 2025) ***(Fig 5)***. A similar study from the M-Mama program pilot in Shinyanga reported a total of 989 completed referrals, out of these, 30.1% were conducted through the standard ambulance referral system, while the majority, 69.9%, utilized the EmTS, highlighting its critical role in bridging transport gaps for maternal emergencies [11].

Mothers were also asked about their reasons for utilizing or not utilizing the M-Mama EmTS during obstetric and neonatal emergencies. Findings revealed that 42% of lactating mothers reported that the emergency circumstances prevented them from using the M-Mama EmTS, while 22% indicated that they were not aware of the service. Conversely, 20% of mothers reported utilizing the M-Mama EmTS because it is a free service, and 16% cited its quick response time as the reason for utilizing ***(Fig 6)***. These findings highlight both demand-side barriers, such as limited awareness and emergency-related constraints, and facilitating factors, such as affordability and timeliness. Similar study on Magnitude and Factors Associated with Ambulance Service Utilization Among Women Who Gave Birth at Public Health Institutions in Central Ethiopia, among the study participants who gave birth at public health institutions but did not utilize the ambulance, several reasons for non-utilization were reported. Some respondents stated that they did not know the ambulance phone number, while others mentioned the lack of ambulance services for returning home after delivery [32]. A study conducted in Jimma City, Ethiopia, on the practice and determinants of ambulance service utilization found that in 105 cases (59.0%), the decision to use an ambulance was made by health workers. In contrast, in 5 cases (2.4%), the decision was made by family members or bystanders. The study also highlighted those perceptions regarding cost and time strongly influenced ambulance use, with many patients opting for ambulances because the service was free and offered shorter waiting times compared to alternative transport options [36].

A cross-tabulation was conducted to assess the utilization of the M-Mama EmTS across different health facilities in Kigoma District Council. Utilization was highest at Chankele Dispensary (93.3%) and Bubango Dispensary (6.7%) among mothers surveyed at these facilities. In contrast, no utilization was reported at Kiganza Dispensary, Kimbwela Dispensary, Mwamgongo Health Center, and Pamila Dispensary during the survey period (0.0%) ***(Fig 7)***. These findings indicate that M-Mama EmTS utilisation varies considerably across facilities, suggesting differences in service awareness, accessibility, or facility-level implementation. This variation may be explained by several factors. Firstly, awareness and knowledge of the M-Mama program are critical [9]; facilities with higher utilization likely benefit from better community sensitization, health education during antenatal care (ANC) visits, and local campaigns [33, 41].

## CONCLUSION

The M-Mama Emergency Transport System (EmTS) utilization among lactating mothers in Kigoma District Council was low but varied uptake across health facilities. Most mothers used emergency transport only once, with private transport remaining the predominant option. Utilization of M-Mama EmTS was strongly influenced by awareness of the service, maternal income, education level, residence, parity, and distance to health facilities.

These findings underscore the need for targeted interventions to improve awareness, ensure operational readiness, and strengthen community engagement across all health facilities to achieve more equitable use of emergency transport services.

## Supporting information

Ethical Approval

Questionnaire Table

Data Set

Stata Do-file for Data Analysis

## Data Availability

The datasets generated and analyses during the current study are available from the corresponding author on reasonable request.

https://ee-eu.kobotoolbox.org/x/1kwKzvN0

## ACKNOWLEDGEMENT

Above all, we give glory and honor to the Almighty God. We would like to acknowledge our beloved colleagues at the Health System department, School of Public Administration and Management, Mzumbe University, Morogoro, Tanzania. Lastly, to everyone who touched or involved in this work in one way or another.

## SUPPORTING INFORMATION

**S1 Data Collection Letters Approval** (DOCX)

**S2 Questionnaire Table** (DOCX)

**S3 Data Set** (XLSX)

**S4 Do File (Do.)**

## Notes

### Competing Interest Statement

The authors have declared no competing interest.

### Funding Statement

The authors received no specific funding for this work.

### Author Declarations

Ethical approval to conduct this evaluation study was obtained from 1. Office of Deputy Vice Chancellor (Academic, Research, and Consultancy), Mzumbe University, Morogoro Region, Tanzania Approval granted, Letter Ref. No. MU/DPGS/INT/38/Vol.IV/592 of March 14, 2025. 2. Office of District Executive Director (DED), Kigoma District Council, Tanzania Approval granted, Letter Ref. No. KDC/H6/1/1/1 of May 13, 2025. 3. Office of District Executive Director (DED), Kigoma District Council, Tanzania, to Health Facilities (Introduction and Permission to conduct data collection from 21 health facilities of Kigoma District Council) Approval granted, Letter Ref. No. KDC/H1/10 of May 13, 2025. 4. Written informed consent was provided to participants before data collection, either by signing or fingerprinting depending on literacy, after being briefed in the local language on the study's purpose, procedures, risks, and benefits. Consent forms were stored separately from the study data, which contained no personal identifiers and was fully anonymised for analysis.

### Summary of Updates

This version of the manuscript has been revised to ensure full compliance with submission requirements. The manuscript title has been aligned with the submission metadata to include the study location and design. The Human Participants Research Checklist has been completed and uploaded. Minor formatting corrections have also been made.

## REFERENCE

[1] World Health Organization (WHO). Maternal Mortality, https://www.who.int/news-room/fact-sheets/detail/maternal-mortality (2025, accessed 8 September 2025).

[2] Geremew AB, Roberts CT, Kassa BG, et al. Exploring evidence of healthcare-seeking pathways for maternal complications in Sub-Saharan Africa: a scoping review. BMC Pregnancy Childbirth; 25. Epub ahead of print 1 December 2025. DOI: 10.1186/s12884-025-07745-3.

[3] Schmitz MM, Serbanescu F, Arnott GE, et al. Referral transit time between sending and first-line receiving health facilities: A geographical analysis in Tanzania. BMJ Glob Health; 4. Epub ahead of print 1 July 2020. DOI: 10.1136/bmjgh-2019-001568.

[4] Thaddeus’ S, Maine D. TOO FAR TO WALK: MATERNAL MORTALITY IN CONTEXT. 1994.

[5] Shah B, Krishnan N, Kodish SR, et al. Applying the Three Delays Model to understand emergency care seeking and delivery in District Bangladesh: A qualitative study. BMJ Open; 10. Epub ahead of print 23 December 2020. DOI: 10.1136/bmjopen-2020-042690.

[6] Actis Danna V, Bedwell C, Wakasiaka S, et al. Utility of the three-delays model and its potential for supporting a solution-based approach to accessing intrapartum care in low- and middle-income countries. A qualitative evidence synthesis. Global Health Action; 13. Epub ahead of print 31 December 2020. DOI: 10.1080/16549716.2020.1819052.

[7] Hadaro TS, Shewangizaw M, Geltore TE, et al. Delays in the decision to seek care and associated factors among women in Ethiopia: systematic review and meta-analysis. BMC Pregnancy Childbirth; 25. Epub ahead of print 1 December 2025. DOI: 10.1186/s12884-025-07968-4.

[8] Pathfinder International. M-Mama: An innovative approach to emergency transportation for mothers and newborns in Tanzania., https://touchfoundation.org/news/the-powerful-impact-of-the-m-mama-emergency-transport-system/ (2021, accessed 8 September 2025).

[9] Sanga A, Kibusi S, Kengia JT. Effectiveness of Community Engagement Using M-Mama Champions in Improving Literacy of Obstetric Danger Signs, Birth Preparedness and Complication Readiness Among Pregnant Women in Bahi, Dodoma. A Community-Based, Cluster Randomized Controlled Trial. Epub ahead of print 2 April 2024. DOI: 10.21203/rs.3.rs-4147830/v1.

[10] Ngoma T, Asiimwe AR, Mukasa J, et al. Addressing the Second Delay in Saving Mothers, Giving Life Districts in Uganda and Zambia: Reaching Appropriate Maternal Care in a Timely Manner, www.ghspjournal.org (2019).

[11] Munishi C, Mateshi G, Mlunde LB, et al. Community-based transport system in Shinyanga, Tanzania: A local innovation averting delays to access health care for maternal emergencies. PLOS Global Public Health; 3. Epub ahead of print 1 August 2023. DOI: 10.1371/journal.pgph.0001487.

[12] Njiro BJ, Ngowi JE, Mlunde L, et al. Towards sustainable emergence transportation system for maternal and new born: Lessons from the m-mama innovative pilot program in Shinyanga, Tanzania. PLOS Global Public Health; 3. Epub ahead of print 1 June 2023. DOI: 10.1371/journal.pgph.0002097.

[13] Berihun A, Abebo TA, Aseffa BM, et al. Third delay and associated factors among women who gave birth at public health facilities of Gurage zone, southern Ethiopia. BMC Womens Health; 23. Epub ahead of print 1 December 2023. DOI: 10.1186/s12905-023-02526-6.

[14] Africa CDC. Tanzania’s Success to Reduce Maternal Mortality Ushers in a Model for Africa, https://africacdc.org/news-item/tanzanias-success-to-reduce-maternal-mortality-ushers-in-a-model-for-africa/ (2025, accessed 8 September 2025).

[15] Tanzania Demographic and Health Survey and Malaria Indicator Survey T-M 2016-2022. Maternal Mortality, https://www.nbs.go.tz/statistics/topic/health-statistics (19 December 2023, accessed 8 September 2025).

[16] Chen YN, Schmitz MM, Serbanescu F, et al. Geographic Access Modeling of Emergency Obstetric and Neonatal Care in Kigoma Region, Tanzania: Transportation Schemes and Programmatic Implications, www.ghspjournal.org (2017).

[17] World Health Organization (WHO). WHO donates Ambulances to reduce maternal deaths in Tanzania. 2023.

[18] Tanzania National Bureau of Statistics (TNBS). Tanzania National Bureau of Statistics (TNBS); Kigoma Region Report, https://www.nbs.go.tz/search?q=Census&button= (2023, accessed 8 September 2025).

[19] Setia MS. Methodology series module 3: Cross-sectional studies. Indian J Dermatol 2016; 61: 261–264.

[20] Creswell JW, & CJD (2018). Research design: Qualitative, quantitative, and mixed methods approaches (5th ed.). SAGE Publications. Fifth edition. Los Angeles: SAGE Publications, Inc., 2018.

[21] Kothari CR. Research methodology: Methods and techniques. Second Edition. New Age International (P) Limited, Publishers., 2004.

[22] McHugh ML. The Chi-square test of independence. Biochem Med (Zagreb) 2013; 23: 143–149.

[23] 1119405262-36.

[24] Anasel MG, Mlinga UJ. Determinants of contraceptive use among married women in Tanzania: Policy implication. Etude de la Population Africaine 2014; 28: 978–988.

[25] Anasel MG, Swai IL. Intimate partner violence, Sexual Violence. Physical/Emotional/Psychological violence 2024; 7: 2024.

[26] Anasel MG, Swai IL. Factors to determine the adoption of online teaching in Tanzania’s Universities during the COVID-19 pandemic. PLoS One; 18. Epub ahead of print 1 October 2023. DOI: 10.1371/journal.pone.0292065.

[27] Banke-Thomas A, Wong KLM, Collins L, et al. An assessment of geographical access and factors influencing travel time to emergency obstetric care in the urban state of Lagos, Nigeria. Health Policy Plan 2021; 36: 1384–1396.

[28] Kim C, Erim D, Natiq K, et al. Combination of Interventions Needed to Improve Maternal Healthcare Utilization: A Multinomial Analysis of the Inequity in Place of Childbirth in Afghanistan. Front Glob Womens Health; 1. Epub ahead of print 2020. DOI: 10.3389/fgwh.2020.571055.

[29] International ethical guidelines for health-related research involving humans. CIOMS, 2017.

[30] Capili B, Anastasi JK. Methods to Disseminate Nursing Research: A Brief Overview Developing a communications strategy in advance is key. American Journal of Nursing 2024; 124: 36–39.

[31] Nunes JMB, Martins JT, Zhou L. more authors) (2010) Contextual Sensitivity in Grounded Theory: The Role of Pilot Studies. The Electronic Journal of Business Research Methods 2010; 8: 73–84.

[32] Tolossa W, Bala ET, Dufera A, et al. Magnitude and Factors Associated with Ambulance Service Utilization Among Women Who Gave Birth at Public Health Institutions in Central Ethiopia. Open Access Emergency Medicine 2022; 14: 457–471.

[33] Wirtu R, Yeshanew S, Geda A. Utilization of Ambulance Services and Associated Factors during Pregnancy and Labor Among Lactating Mothers in Buno Bedele Zone, Southwest Ethiopia. Health Serv Insights; 16. Epub ahead of print 1 January 2023. DOI: 10.1177/11786329231157227.

[34] Langa N, Bhatta T. The District-urban divide in Tanzania: Residential context and socioeconomic inequalities in maternal health care utilization. PLoS One; 15. Epub ahead of print 1 November 2020. DOI: 10.1371/journal.pone.0241746.

[35] Banke-Thomas A, Beňová L, Ray N, et al. Inequalities in geographical access to emergency obstetric and newborn care.

[36] Adem MA, Tezera ZB, Agegnehu CD. The practice and determinants of ambulance service utilization in pre-hospital settings, Jimma City, Ethiopia. BMC Emerg Med; 24. Epub ahead of print 1 December 2024. DOI: 10.1186/s12873-024-00999-8.

[37] Xu X, Zhang Q, You H, et al. Awareness, Utilization and Health Outcomes of National Essential Public Health Service Among Migrants in China. Front Public Health; 10. Epub ahead of print 11 July 2022. DOI: 10.3389/fpubh.2022.936275.

[38] Leo GM, Ekele EP, Chiejina EE, et al. Awareness, Perceptions and Utilization of Cervical Screening Services among Women of Child – Bearing Age in Abuja, Nigeria. Texila International Journal of Public Health; 8. Epub ahead of print 10 September 2020. DOI: 10.21522/TIJPH.2013.08.03.

[39] Li Y, Li H, Jiang Y. Factors influencing maternal healthcare utilization in Papua New Guinea: Andersen’s behaviour model. BMC Womens Health; 23. Epub ahead of print 1 December 2023. DOI: 10.1186/s12905-023-02709-1.

[40] Shanto HH, Al-Zubayer MA, Ahammed B, et al. Maternal Healthcare Services Utilisation and Its Associated Risk Factors: A Pooled Study of 37 Low- and Middle-Income Countries. Int J Public Health; 68. Epub ahead of print 2023. DOI: 10.3389/ijph.2023.1606288.

[41] Okedo-Alex IN, Akamike IC, Ezeanosike OB, et al. Determinants of antenatal care utilisation in sub-Saharan Africa: A systematic review. BMJ Open; 9. Epub ahead of print 1 October 2019. DOI: 10.1136/bmjopen-2019-031890.

[42] Furechi O. Awareness and utilization of free maternal healthcare services among women in Mt. Elgon Sub-County, Kenya. Epub ahead of print 28 January 2023. DOI: 10.1101/2023.01.25.23285004.

[43] Framework on integrated, people-centred health services Report by the Secretariat, http://apps.who.int/iris/bitstream/10665/174536/1/9789241564977_eng.pdf?ua=1, (2016).

[44] Abebe TA, Debelew GT. Maternal health care services utilization and associated factors among pregnant women in Kersa district, Jimma zone, Southwest Ethiopia. PLoS One; 20. Epub ahead of print 1 May 2025. DOI: 10.1371/journal.pone.0323977.

[45] Harring AKV, Graesli O, Häikiö K, et al. Frequent contacts to Emergency Medical Services (EMS): more than frequent callers. BMC Emerg Med; 24. Epub ahead of print 1 December 2024. DOI: 10.1186/s12873-024-01104-9.

[46] Adam VY, Awunor NS. PERCEPTIONS AND FACTORS AFFECTING UTILIZATION OF HEALTH SERVICES IN A DISTRICT COMMUNITY IN SOUTHERN NIGERIA 1 *2*. 2014.

[47] Orok E, Nwifama S, Oni O, et al. Students’ perception of healthcare services and factors affecting their utilization at a Nigerian University: a cross-sectional study. Sci Rep; 14. Epub ahead of print 1 December 2024. DOI: 10.1038/s41598-024-75573-0.

[48] Shayo EH, Nassor NK, Mboera LEG, et al. The impacts of COVID-19 and its policy response on access and utilization of maternal and child health services in Tanzania: A mixed methods study. PLOS Global Public Health; 3. Epub ahead of print 1 May 2023. DOI: 10.1371/journal.pgph.0001549.

