## Supplementary material for "Outcome evaluation of the M-Mama emergency transport system (EmTS) and factors associated with its utilisation among lactating mothers in Kigoma District Council, Tanzania: A cross-sectional study": Ethical Approval

UNITED REPUBLIC OF TANZANIA
MINISTRY OF EDUCATION, SCIENCE AND TECHNOLOGY

MZUMBE UNIVERSITY
OFFICE OF THE DEPUTY VICE CHANCELLOR
(ARC)


Website: www.mzumbe.ac.tz

P.O. BOX 1
MZUMBE
MOROGORO, TANZANIA

*In replying please quote:*

**Ref. No. MU/DPGS/INT/38/Vol. IV/592** **14th March, 2025**

Kigoma District Council,
P.O.BOX 555
**47204 KIGOMA.**

**RE: INTRODUCTION OF MR. JULIUS MASABA**

Refer to the heading captioned above

2. The bearer of this letter **Mr. Julius Masaba** whose registration number is 1842259/T.23 is a postgraduate student at our University (Mzumbe University) pursuing **Master of Science in Health Monitoring and Evaluation (MSc.HM&E)**. As part of requirements for completion of his studies, he is collecting data on: "*Outcome Evaluation of the M-Mama Emergency Transport System (EmTS) and factors associated with its utilisation among lactating Mothers in Kigoma District Council*". The activity is expected to start at 1st May up to 30rd August 2025.

3. This letter serves to achieve three purposes. Firstly, to introduce him to you. Secondly, to request you to grant him permission to undertake data collection at your organizations, and thirdly to request you to facilitate any form of assistance he might need in order to successfully pursue this noble exercise at your organization. We can assure you that this activity is entirely for academic purpose and will never be used for any other purposes.

4. We trust that you will accord our student with necessary assistance.

5. Sincerely yours,

MZUMBE UNIVERSITY
P. O. Box 1, MZUMBE
TANZANIA

Dr. Juma Buhimila (PhD)

**FOR DEPUTY VICE CHANCELLOR (Academic, Research and Consultancy)**

**THE UNITED REPUBLIC OF TANZANIA**
**PRESIDENT'S OFFICE**

**REGIONAL GOVERNMENTS AND LOCAL GOVERNMENTS**

**KIGOMA DISTRICT COUNCIL**


Tovuti: www.kigomadc.go.tz

Executive Director's Office
P. O Box 332,
**KIGOMA.**

**REF.NO.KDC/H6/1/1/1** May 13, 2025.

Mr. Julius Masaba

Mzumbe University,

P.O. Box 1,

Morogoro,

**TANZANIA**

**RE: APPROVAL TO CONDUCT DATA COLLECTION ON OUTCOME EVALUATION OF THE M-MAMA EMERGENCY TRANSPORT SYSTEM (EMTS) AND FACTORS ASSOCIATED WITH ITS UTILISATION AMONG LACTATING MOTHERS IN KIGOMA DISTRICT COUNCIL**

Please refer to the subject mentioned above.

2. Reference is made to your request with **Ref. No. Mu/DPGS/INT/38/Vol.IV/592** from Mzumbe University regarding permission to conduct data collection for academic purposes in health facilities located within Kigoma District Council.

3. With this letter we're pleased to inform you that your request has been approved. You are therefore authorized to proceed with the data collection process in the health facilities under Kigoma District Council from the date of this letter to 30rd August, 2025.

4. Please ensure that all ethical considerations and research protocols are strictly followed throughout your data collection activities.

5. We wish you all the best in your research.

..................

DISTRICT EXECUTIVE DIRECTOR
P.O.BOX 332
KIGOMA

Chiriku H. Chilumba
**EXECUTIVE DIRECTOR**
**KIGOMA DISTRICT COUNCIL**

**CC:** Deputy Vice Chancellor (Academic, Research and Consultancy)
Mzumbe University
P. O Box 1
Morogoro, Tanzania

**THE UNITED REPUBLIC OF TANZANIA**
**PRESIDENT'S OFFICE**
**REGIONAL GOVERNMENTS AND LOCAL GOVERNMENTS**
**KIGOMA DISTRICT COUNCIL**

KIGOMA DISTRICT COUNCIL
P. O. Box 332


Tovuti: www.kigomadc.go.tz

Executive Director's Office
P. O Box 332,
**KIGOMA.**

---

**REF. NO: KDC/H/1/10**

May 13, 2025.

Medical Officer In-Charge,
Bitale Health Centre,
P.O. Box 555,
**KIGOMA.**

**RE: INTRODUCTION AND PERMISSION FOR DATA COLLECTION ON OUTCOME EVALUATION OF THE M-MAMA EMERGENCY TRANSPORT SYSTEM (EMTS) AND FACTORS ASSOCIATED WITH ITS UTILISATION AMONG LACTATING MOTHERS IN KIGOMA DISTRICT COUNCIL**

Please refer to the subject mentioned above.

2. Through this letter, I would like to introduce to you **Mr. Julius Masaba** from Mzumbe University, who is conducting a study titled: "*Outcome evaluation of the M-Mama emergency transport system (EmTS) and factors associated with its utilisation among lactating mothers in Kigoma District Council.*"

3. The purpose of this study is to examine the factors associated with the use of the M-Mama emergency transport system in improving the health of mothers and newborns. The researcher will collect data from women who delivered at the health facility and from those attending the MCH clinic at your facility, starting from the date of this letter until 30rd August 2025.

4. Kindly provide full cooperation to this researcher. Also, instruct the staff to offer all necessary assistance to facilitate the smooth completion of the study.

5. I wish you successful implementation.

..................

DISTRICT EXECUTIVE DIRECTOR
P.O.BOX 332
KIGOMA

Chiriku H. Chilumba
**EXECUTIVE DIRECTOR**
**KIGOMA DISTRICT COUNCIL**

**CC:** Deputy Vice Chancellor (Academic, Research and Consultancy)
Mzumbe University
P. O Box 1
Morogoro, Tanzania

**THE UNITED REPUBLIC OF TANZANIA**
**PRESIDENT'S OFFICE**
**REGIONAL GOVERNMENTS AND LOCAL GOVERNMENTS**
**KIGOMA DISTRICT COUNCIL**


Tovuti: www.kigomadc.go.tz

Executive Director's Office
P. O Box 332,
**KIGOMA.**

---

**REF. NO: KDC/H/1/10**

May 13, 2025.

Medical Officer In-Charge,
Bubango Dispensary,
P.O. Box 555,
**KIGOMA.**

**RE: INTRODUCTION AND PERMISSION FOR DATA COLLECTION ON OUTCOME EVALUATION OF THE M-MAMA EMERGENCY TRANSPORT SYSTEM (EMTS) AND FACTORS ASSOCIATED WITH ITS UTILISATION AMONG LACTATING MOTHERS IN KIGOMA DISTRICT COUNCIL**

Please refer to the subject mentioned above.

2. Through this letter, I would like to introduce to you **Mr. Julius Masaba** from Mzumbe University, who is conducting a study titled: "*Outcome evaluation of the M-Mama emergency transport system (EmTS) and factors associated with its utilisation among lactating mothers in Kigoma District Council.*"

3. The purpose of this study is to examine the factors associated with the use of the M-Mama emergency transport system in improving the health of mothers and newborns. The researcher will collect data from women who delivered at the health facility and from those attending the MCH clinic at your facility, starting from the date of this letter until 30rd August 2025.

4. Kindly provide full cooperation to this researcher. Also, instruct the staff to offer all necessary assistance to facilitate the smooth completion of the study.

5. I wish you successful implementation.

DISTRICT EXECUTIVE DIRECTOR
P.O.BOX 332
KIGOMA

………………
Chiriku H. Chilumba
**EXECUTIVE DIRECTOR**
**KIGOMA DISTRICT COUNCIL**

**CC:** Deputy Vice Chancellor (Academic, Research and Consultancy)
Mzumbe University
P. O Box 1
Morogoro, Tanzania

**THE UNITED REPUBLIC OF TANZANIA**
**PRESIDENT'S OFFICE**
**REGIONAL GOVERNMENTS AND LOCAL GOVERNMENTS**
**KIGOMA DISTRICT COUNCIL**


Tovuti: www.kigomadc.go.tz

Executive Director's Office
P. O Box 332,
**KIGOMA.**

---

**REF. NO: KDC/H/1/10**

May 13, 2025.

Medical Officer In-Charge,
Chankabwimba Dispensary,
P.O. Box 555,
**KIGOMA.**

**RE: INTRODUCTION AND PERMISSION FOR DATA COLLECTION ON OUTCOME EVALUATION OF THE M-MAMA EMERGENCY TRANSPORT SYSTEM (EMTS) AND FACTORS ASSOCIATED WITH ITS UTILISATION AMONG LACTATING MOTHERS IN KIGOMA DISTRICT COUNCIL**

Please refer to the subject mentioned above.

2. Through this letter, I would like to introduce to you **Mr. Julius Masaba** from Mzumbe University, who is conducting a study titled: "*Outcome evaluation of the M-Mama emergency transport system (EmTS) and factors associated with its utilisation among lactating mothers in Kigoma District Council.*"

3. The purpose of this study is to examine the factors associated with the use of the M-Mama emergency transport system in improving the health of mothers and newborns. The researcher will collect data from women who delivered at the health facility and from those attending the MCH clinic at your facility, starting from the date of this letter until 30rd August 2025.

4. Kindly provide full cooperation to this researcher. Also, instruct the staff to offer all necessary assistance to facilitate the smooth completion of the study.

5. I wish you successful implementation.

..................

DISTRICT EXECUTIVE DIRECTOR
P.O.BOX 332
KIGOMA

Chiriku H. Chilumba
**EXECUTIVE DIRECTOR**
**KIGOMA DISTRICT COUNCIL**

**CC:** Deputy Vice Chancellor (Academic, Research and Consultancy)
Mzumbe University
P. O Box 1
Morogoro, Tanzania

**THE UNITED REPUBLIC OF TANZANIA**
**PRESIDENT'S OFFICE**
**REGIONAL GOVERNMENTS AND LOCAL GOVERNMENTS**
**KIGOMA DISTRICT COUNCIL**


Tovuti: www.kigomadc.go.tz

Executive Director's Office
P. O Box 332,
**KIGOMA.**

---

**REF. NO: KDC/H/1/10**

May 13, 2025.

Medical Officer In-Charge,
Kagongo Dispensary,
P.O. Box 555,
**KIGOMA.**

**RE: INTRODUCTION AND PERMISSION FOR DATA COLLECTION ON OUTCOME EVALUATION OF THE M-MAMA EMERGENCY TRANSPORT SYSTEM (EMTS) AND FACTORS ASSOCIATED WITH ITS UTILISATION AMONG LACTATING MOTHERS IN KIGOMA DISTRICT COUNCIL**

Please refer to the subject mentioned above.

2. Through this letter, I would like to introduce to you **Mr. Julius Masaba** from Mzumbe University, who is conducting a study titled: "*Outcome evaluation of the M-Mama emergency transport system (EmTS) and factors associated with its utilisation among lactating mothers in Kigoma District Council.*"

3. The purpose of this study is to examine the factors associated with the use of the M-Mama emergency transport system in improving the health of mothers and newborns. The researcher will collect data from women who delivered at the health facility and from those attending the MCH clinic at your facility, starting from the date of this letter until 30rd August 2025.

4. Kindly provide full cooperation to this researcher. Also, instruct the staff to offer all necessary assistance to facilitate the smooth completion of the study.

5. I wish you successful implementation.

..................
Chiriku H. Chilumba
**EXECUTIVE DIRECTOR**
**KIGOMA DISTRICT COUNCIL**

DISTRICT EXECUTIVE DIRECTOR
P.O.BOX 332
KIGOMA

**CC:** Deputy Vice Chancellor (Academic, Research and Consultancy)
Mzumbe University
P. O Box 1
Morogoro, Tanzania

**THE UNITED REPUBLIC OF TANZANIA**
**PRESIDENT'S OFFICE**
**REGIONAL GOVERNMENTS AND LOCAL GOVERNMENTS**
**KIGOMA DISTRICT COUNCIL**


Tovuti: www.kigomadc.go.tz

Executive Director's Office
P. O Box 332,
**KIGOMA.**

**REF. NO: KDC/H/1/10** May 13, 2025.

Medical Officer In-Charge,
Kagunga Dispensary,
P.O. Box 555,
**KIGOMA.**

**RE: INTRODUCTION AND PERMISSION FOR DATA COLLECTION ON OUTCOME EVALUATION OF THE M-MAMA EMERGENCY TRANSPORT SYSTEM (EMTS) AND FACTORS ASSOCIATED WITH ITS UTILISATION AMONG LACTATING MOTHERS IN KIGOMA DISTRICT COUNCIL**

Please refer to the subject mentioned above.

2. Through this letter, I would like to introduce to you **Mr. Julius Masaba** from Mzumbe University, who is conducting a study titled: "*Outcome evaluation of the M-Mama emergency transport system (EmTS) and factors associated with its utilisation among lactating mothers in Kigoma District Council.*"

3. The purpose of this study is to examine the factors associated with the use of the M-Mama emergency transport system in improving the health of mothers and newborns. The researcher will collect data from women who delivered at the health facility and from those attending the MCH clinic at your facility, starting from the date of this letter until 30rd August 2025.

4. Kindly provide full cooperation to this researcher. Also, instruct the staff to offer all necessary assistance to facilitate the smooth completion of the study.

5. I wish you successful implementation.

..................
Chiriku H. Chilumba
**EXECUTIVE DIRECTOR**
**KIGOMA DISTRICT COUNCIL**

DISTRICT EXECUTIVE DIRECTOR
P.O.BOX 332
KIGOMA

**CC:** Deputy Vice Chancellor (Academic, Research and Consultancy)
Mzumbe University
P. O Box 1
Morogoro, Tanzania

**THE UNITED REPUBLIC OF TANZANIA**
**PRESIDENT'S OFFICE**
**REGIONAL GOVERNMENTS AND LOCAL GOVERNMENTS**
**KIGOMA DISTRICT COUNCIL**


Tovuti: www.kigomadc.go.tz

Executive Director's Office
P. O Box 332,
**KIGOMA.**

---

**REF. NO: KDC/H/1/10**

May 13, 2025.

Medical Officer In-Charge,
Kalalangabo Dispensary,
P.O. Box 555,
**KIGOMA.**

**RE: INTRODUCTION AND PERMISSION FOR DATA COLLECTION ON OUTCOME EVALUATION OF THE M-MAMA EMERGENCY TRANSPORT SYSTEM (EMTS) AND FACTORS ASSOCIATED WITH ITS UTILISATION AMONG LACTATING MOTHERS IN KIGOMA DISTRICT COUNCIL**

Please refer to the subject mentioned above.

2. Through this letter, I would like to introduce to you **Mr. Julius Masaba** from Mzumbe University, who is conducting a study titled: "*Outcome evaluation of the M-Mama emergency transport system (EmTS) and factors associated with its utilisation among lactating mothers in Kigoma District Council.*"

3. The purpose of this study is to examine the factors associated with the use of the M-Mama emergency transport system in improving the health of mothers and newborns. The researcher will collect data from women who delivered at the health facility and from those attending the MCH clinic at your facility, starting from the date of this letter until 30rd August 2025.

4. Kindly provide full cooperation to this researcher. Also, instruct the staff to offer all necessary assistance to facilitate the smooth completion of the study.

5. I wish you successful implementation.

DISTRICT EXECUTIVE DIRECTOR
P.O.BOX 332
KIGOMA

………………
Chiriku H. Chilumba
**EXECUTIVE DIRECTOR**
**KIGOMA DISTRICT COUNCIL**

**CC:** Deputy Vice Chancellor (Academic, Research and Consultancy)
Mzumbe University
P. O Box 1
Morogoro, Tanzania

**THE UNITED REPUBLIC OF TANZANIA**
**PRESIDENT’S OFFICE**
**REGIONAL GOVERNMENTS AND LOCAL GOVERNMENTS**
**KIGOMA DISTRICT COUNCIL**


Tovuti: www.kigomadc.go.tz

Executive Director’s Office
P. O Box 332,
**KIGOMA.**

**REF. NO: KDC/H/1/10** May 13, 2025.

Medical Officer In-Charge,
Kamara Dispensary,
P.O. Box 555,
**KIGOMA.**

**RE: INTRODUCTION AND PERMISSION FOR DATA COLLECTION ON OUTCOME EVALUATION OF THE M-MAMA EMERGENCY TRANSPORT SYSTEM (EMTS) AND FACTORS ASSOCIATED WITH ITS UTILISATION AMONG LACTATING MOTHERS IN KIGOMA DISTRICT COUNCIL**

Please refer to the subject mentioned above.

2. Through this letter, I would like to introduce to you **Mr. Julius Masaba** from Mzumbe University, who is conducting a study titled: “*Outcome evaluation of the M-Mama emergency transport system (EmTS) and factors associated with its utilisation among lactating mothers in Kigoma District Council.*”

3. The purpose of this study is to examine the factors associated with the use of the M-Mama emergency transport system in improving the health of mothers and newborns. The researcher will collect data from women who delivered at the health facility and from those attending the MCH clinic at your facility, starting from the date of this letter until 30rd August 2025.

4. Kindly provide full cooperation to this researcher. Also, instruct the staff to offer all necessary assistance to facilitate the smooth completion of the study.

5. I wish you successful implementation.

……………….

Chiriku H. Chilumba
**EXECUTIVE DIRECTOR**
**KIGOMA DISTRICT COUNCIL**

DISTRICT EXECUTIVE DIRECTOR
P.O.BOX 332
KIGOMA

**CC:** Deputy Vice Chancellor (Academic, Research and Consultancy)
Mzumbe University
P. O Box 1
Morogoro, Tanzania

**THE UNITED REPUBLIC OF TANZANIA**
**PRESIDENT'S OFFICE**
**REGIONAL GOVERNMENTS AND LOCAL GOVERNMENTS**
**KIGOMA DISTRICT COUNCIL**


Tovuti: www.kigomadc.go.tz

Executive Director's Office
P. O Box 332,
**KIGOMA.**

---

**REF. NO: KDC/H/1/10**

May 13, 2025.

Medical Officer In-Charge,
Kaseke Dispensary,
P.O. Box 555,
**KIGOMA.**

**RE: INTRODUCTION AND PERMISSION FOR DATA COLLECTION ON OUTCOME EVALUATION OF THE M-MAMA EMERGENCY TRANSPORT SYSTEM (EMTS) AND FACTORS ASSOCIATED WITH ITS UTILISATION AMONG LACTATING MOTHERS IN KIGOMA DISTRICT COUNCIL**

Please refer to the subject mentioned above.

2. Through this letter, I would like to introduce to you **Mr. Julius Masaba** from Mzumbe University, who is conducting a study titled: "*Outcome evaluation of the M-Mama emergency transport system (EmTS) and factors associated with its utilisation among lactating mothers in Kigoma District Council.*"

3. The purpose of this study is to examine the factors associated with the use of the M-Mama emergency transport system in improving the health of mothers and newborns. The researcher will collect data from women who delivered at the health facility and from those attending the MCH clinic at your facility, starting from the date of this letter until 30rd August 2025.

4. Kindly provide full cooperation to this researcher. Also, instruct the staff to offer all necessary assistance to facilitate the smooth completion of the study.

5. I wish you successful implementation.

..................

Chiriku H. Chilumba
**EXECUTIVE DIRECTOR**
**KIGOMA DISTRICT COUNCIL**

DISTRICT EXECUTIVE DIRECTOR
P.O.BOX 332
KIGOMA

**CC:** Deputy Vice Chancellor (Academic, Research and Consultancy)
Mzumbe University
P. O Box 1
Morogoro, Tanzania

**THE UNITED REPUBLIC OF TANZANIA**
**PRESIDENT'S OFFICE**
**REGIONAL GOVERNMENTS AND LOCAL GOVERNMENTS**
**KIGOMA DISTRICT COUNCIL**


Tovuti: www.kigomadc.go.tz

Executive Director's Office
P. O Box 332,
**KIGOMA.**

---

**REF. NO: KDC/H/1/10**

May 13, 2025.

Medical Officer In-Charge,
Kiganza Dispensary,
P.O. Box 555,
**KIGOMA.**

**RE: INTRODUCTION AND PERMISSION FOR DATA COLLECTION ON OUTCOME EVALUATION OF THE M-MAMA EMERGENCY TRANSPORT SYSTEM (EMTS) AND FACTORS ASSOCIATED WITH ITS UTILISATION AMONG LACTATING MOTHERS IN KIGOMA DISTRICT COUNCIL**

Please refer to the subject mentioned above.

2. Through this letter, I would like to introduce to you **Mr. Julius Masaba** from Mzumbe University, who is conducting a study titled: "*Outcome evaluation of the M-Mama emergency transport system (EmTS) and factors associated with its utilisation among lactating mothers in Kigoma District Council.*"

3. The purpose of this study is to examine the factors associated with the use of the M-Mama emergency transport system in improving the health of mothers and newborns. The researcher will collect data from women who delivered at the health facility and from those attending the MCH clinic at your facility, starting from the date of this letter until 30rd August 2025.

4. Kindly provide full cooperation to this researcher. Also, instruct the staff to offer all necessary assistance to facilitate the smooth completion of the study.

5. I wish you successful implementation.

……………….

DISTRICT EXECUTIVE DIRECTOR
P.O.BOX 332
KIGOMA

Chiriku H. Chilumba
**EXECUTIVE DIRECTOR**
**KIGOMA DISTRICT COUNCIL**

**CC:** Deputy Vice Chancellor (Academic, Research and Consultancy)
Mzumbe University
P. O Box 1
Morogoro, Tanzania

**THE UNITED REPUBLIC OF TANZANIA**
**PRESIDENT'S OFFICE**
**REGIONAL GOVERNMENTS AND LOCAL GOVERNMENTS**
**KIGOMA DISTRICT COUNCIL**



Tovuti: www.kigomadc.go.tz

Executive Director's Office
P. O Box 332,
**KIGOMA.**


**REF. NO: KDC/H/1/10** May 13, 2025.

Medical Officer In-Charge,
Kimbwela Dispensary,
P.O. Box 555,
**KIGOMA.**

**RE: INTRODUCTION AND PERMISSION FOR DATA COLLECTION ON OUTCOME EVALUATION OF THE M-MAMA EMERGENCY TRANSPORT SYSTEM (EMTS) AND FACTORS ASSOCIATED WITH ITS UTILISATION AMONG LACTATING MOTHERS IN KIGOMA DISTRICT COUNCIL**

Please refer to the subject mentioned above.

2. Through this letter, I would like to introduce to you **Mr. Julius Masaba** from Mzumbe University, who is conducting a study titled: "*Outcome evaluation of the M-Mama emergency transport system (EmTS) and factors associated with its utilisation among lactating mothers in Kigoma District Council.*"

3. The purpose of this study is to examine the factors associated with the use of the M-Mama emergency transport system in improving the health of mothers and newborns. The researcher will collect data from women who delivered at the health facility and from those attending the MCH clinic at your facility, starting from the date of this letter until 30$^{rd}$ August 2025.

4. Kindly provide full cooperation to this researcher. Also, instruct the staff to offer all necessary assistance to facilitate the smooth completion of the study.

5. I wish you successful implementation.

DISTRICT EXECUTIVE DIRECTOR
P.O.BOX 332
KIGOMA

………………
Chiriku H. Chilumba
**EXECUTIVE DIRECTOR**
**KIGOMA DISTRICT COUNCIL**

**CC:** Deputy Vice Chancellor (Academic, Research and Consultancy)
Mzumbe University
P. O Box 1
Morogoro, Tanzania

**THE UNITED REPUBLIC OF TANZANIA**
**PRESIDENT'S OFFICE**
**REGIONAL GOVERNMENTS AND LOCAL GOVERNMENTS**
**KIGOMA DISTRICT COUNCIL**


Tovuti: www.kigomadc.go.tz

Executive Director's Office
P. O Box 332,
**KIGOMA.**

**REF. NO: KDC/H/1/10**

May 13, 2025.

Medical Officer In-Charge,
Mahembe Dispensary,
P.O. Box 555,
**KIGOMA.**

**RE: INTRODUCTION AND PERMISSION FOR DATA COLLECTION ON OUTCOME EVALUATION OF THE M-MAMA EMERGENCY TRANSPORT SYSTEM (EMTS) AND FACTORS ASSOCIATED WITH ITS UTILISATION AMONG LACTATING MOTHERS IN KIGOMA DISTRICT COUNCIL**

Please refer to the subject mentioned above.

2. Through this letter, I would like to introduce to you **Mr. Julius Masaba** from Mzumbe University, who is conducting a study titled: "*Outcome evaluation of the M-Mama emergency transport system (EmTS) and factors associated with its utilisation among lactating mothers in Kigoma District Council.*"

3. The purpose of this study is to examine the factors associated with the use of the M-Mama emergency transport system in improving the health of mothers and newborns. The researcher will collect data from women who delivered at the health facility and from those attending the MCH clinic at your facility, starting from the date of this letter until 30rd August 2025.

4. Kindly provide full cooperation to this researcher. Also, instruct the staff to offer all necessary assistance to facilitate the smooth completion of the study.

5. I wish you successful implementation.

………………
Chiriku H. Chilumba
**EXECUTIVE DIRECTOR**
**KIGOMA DISTRICT COUNCIL**

DISTRICT EXECUTIVE DIRECTOR
P.O.BOX 332
KIGOMA

**CC:** Deputy Vice Chancellor (Academic, Research and Consultancy)
Mzumbe University
P. O Box 1
Morogoro, Tanzania

**THE UNITED REPUBLIC OF TANZANIA**
**PRESIDENT'S OFFICE**
**REGIONAL GOVERNMENTS AND LOCAL GOVERNMENTS**
**KIGOMA DISTRICT COUNCIL**


Tovuti: www.kigomadc.go.tz

Executive Director's Office
P. O Box 332,
**KIGOMA.**

**REF. NO: KDC/H/1/10** May 13, 2025.

Medical Officer In-Charge,
Matendo Dispensary,
P.O. Box 555,
**KIGOMA.**

**RE: INTRODUCTION AND PERMISSION FOR DATA COLLECTION ON OUTCOME EVALUATION OF THE M-MAMA EMERGENCY TRANSPORT SYSTEM (EMTS) AND FACTORS ASSOCIATED WITH ITS UTILISATION AMONG LACTATING MOTHERS IN KIGOMA DISTRICT COUNCIL**

Please refer to the subject mentioned above.

2. Through this letter, I would like to introduce to you **Mr. Julius Masaba** from Mzumbe University, who is conducting a study titled: "*Outcome evaluation of the M-Mama emergency transport system (EmTS) and factors associated with its utilisation among lactating mothers in Kigoma District Council.*"

3. The purpose of this study is to examine the factors associated with the use of the M-Mama emergency transport system in improving the health of mothers and newborns. The researcher will collect data from women who delivered at the health facility and from those attending the MCH clinic at your facility, starting from the date of this letter until 30rd August 2025.

4. Kindly provide full cooperation to this researcher. Also, instruct the staff to offer all necessary assistance to facilitate the smooth completion of the study.

5. I wish you successful implementation.

..................
Chiriku H. Chilumba
**EXECUTIVE DIRECTOR**
**KIGOMA DISTRICT COUNCIL**

DISTRICT EXECUTIVE DIRECTOR
P.O.BOX 332
KIGOMA

**CC:** Deputy Vice Chancellor (Academic, Research and Consultancy)
Mzumbe University
P. O Box 1
Morogoro, Tanzania

**THE UNITED REPUBLIC OF TANZANIA**
**PRESIDENT'S OFFICE**
**REGIONAL GOVERNMENTS AND LOCAL GOVERNMENTS**
**KIGOMA DISTRICT COUNCIL**


Tovuti: www.kigomadc.go.tz

Executive Director's Office
P. O Box 332,
**KIGOMA.**

**REF. NO: KDC/H/1/10**

May 13, 2025.

Medical Officer In-Charge,
Matyazo Health Center,
P.O. Box 555,
**KIGOMA.**

**RE: INTRODUCTION AND PERMISSION FOR DATA COLLECTION ON OUTCOME EVALUATION OF THE M-MAMA EMERGENCY TRANSPORT SYSTEM (EMTS) AND FACTORS ASSOCIATED WITH ITS UTILISATION AMONG LACTATING MOTHERS IN KIGOMA DISTRICT COUNCIL**

Please refer to the subject mentioned above.

2. Through this letter, I would like to introduce to you **Mr. Julius Masaba** from Mzumbe University, who is conducting a study titled: "*Outcome evaluation of the M-Mama emergency transport system (EmTS) and factors associated with its utilisation among lactating mothers in Kigoma District Council.*"

3. The purpose of this study is to examine the factors associated with the use of the M-Mama emergency transport system in improving the health of mothers and newborns. The researcher will collect data from women who delivered at the health facility and from those attending the MCH clinic at your facility, starting from the date of this letter until 30rd August 2025.

4. Kindly provide full cooperation to this researcher. Also, instruct the staff to offer all necessary assistance to facilitate the smooth completion of the study.

5. I wish you successful implementation.

DISTRICT EXECUTIVE DIRECTOR
P.O.BOX 332
KIGOMA

Chiriku H. Chilumba
**EXECUTIVE DIRECTOR**
**KIGOMA DISTRICT COUNCIL**

**CC:** Deputy Vice Chancellor (Academic, Research and Consultancy)
Mzumbe University
P. O Box 1
Morogoro, Tanzania

**THE UNITED REPUBLIC OF TANZANIA**
**PRESIDENT'S OFFICE**
**REGIONAL GOVERNMENTS AND LOCAL GOVERNMENTS**
**KIGOMA DISTRICT COUNCIL**


Tovuti: www.kigomadc.go.tz

Executive Director's Office
P. O Box 332,
**KIGOMA.**

**REF. NO: KDC/H/1/10** May 13, 2025.

Medical Officer In-Charge,
Mgaraganza Dispensary,
P.O. Box 555,
**KIGOMA.**

**RE: INTRODUCTION AND PERMISSION FOR DATA COLLECTION ON OUTCOME EVALUATION OF THE M-MAMA EMERGENCY TRANSPORT SYSTEM (EMTS) AND FACTORS ASSOCIATED WITH ITS UTILISATION AMONG LACTATING MOTHERS IN KIGOMA DISTRICT COUNCIL**

Please refer to the subject mentioned above.

2. Through this letter, I would like to introduce to you **Mr. Julius Masaba** from Mzumbe University, who is conducting a study titled: "*Outcome evaluation of the M-Mama emergency transport system (EmTS) and factors associated with its utilisation among lactating mothers in Kigoma District Council.*"

3. The purpose of this study is to examine the factors associated with the use of the M-Mama emergency transport system in improving the health of mothers and newborns. The researcher will collect data from women who delivered at the health facility and from those attending the MCH clinic at your facility, starting from the date of this letter until 30rd August 2025.

4. Kindly provide full cooperation to this researcher. Also, instruct the staff to offer all necessary assistance to facilitate the smooth completion of the study.

5. I wish you successful implementation.

..................

Chiriku H. Chilumba
**EXECUTIVE DIRECTOR**
**KIGOMA DISTRICT COUNCIL**

DISTRICT EXECUTIVE DIRECTOR
P.O.BOX 332
KIGOMA

**CC:** Deputy Vice Chancellor (Academic, Research and Consultancy)
Mzumbe University
P. O Box 1
Morogoro, Tanzania

**THE UNITED REPUBLIC OF TANZANIA**
**PRESIDENT'S OFFICE**
**REGIONAL GOVERNMENTS AND LOCAL GOVERNMENTS**
**KIGOMA DISTRICT COUNCIL**


Tovuti: www.kigomadc.go.tz

Executive Director's Office
P. O Box 332,
**KIGOMA.**

---

**REF. NO: KDC/H/1/10**

May 13, 2025.

Medical Officer In-Charge,
Mkongoro Dispensary,
P.O. Box 555,
**KIGOMA.**

**RE: INTRODUCTION AND PERMISSION FOR DATA COLLECTION ON OUTCOME EVALUATION OF THE M-MAMA EMERGENCY TRANSPORT SYSTEM (EMTS) AND FACTORS ASSOCIATED WITH ITS UTILISATION AMONG LACTATING MOTHERS IN KIGOMA DISTRICT COUNCIL**

Please refer to the subject mentioned above.

2. Through this letter, I would like to introduce to you **Mr. Julius Masaba** from Mzumbe University, who is conducting a study titled: "*Outcome evaluation of the M-Mama emergency transport system (EmTS) and factors associated with its utilisation among lactating mothers in Kigoma District Council.*"

3. The purpose of this study is to examine the factors associated with the use of the M-Mama emergency transport system in improving the health of mothers and newborns. The researcher will collect data from women who delivered at the health facility and from those attending the MCH clinic at your facility, starting from the date of this letter until 30rd August 2025.

4. Kindly provide full cooperation to this researcher. Also, instruct the staff to offer all necessary assistance to facilitate the smooth completion of the study.

5. I wish you successful implementation.

………………

DISTRICT EXECUTIVE DIRECTOR
P.O.BOX 332
KIGOMA

Chiriku H. Chilumba
**EXECUTIVE DIRECTOR**
**KIGOMA DISTRICT COUNCIL**

**CC:** Deputy Vice Chancellor (Academic, Research and Consultancy)
Mzumbe University
P. O Box 1
Morogoro, Tanzania

**THE UNITED REPUBLIC OF TANZANIA**
**PRESIDENT'S OFFICE**
**REGIONAL GOVERNMENTS AND LOCAL GOVERNMENTS**
**KIGOMA DISTRICT COUNCIL**

KIGOMA DISTRICT COUNCIL
P. O. Box 332


Tovuti: www.kigomadc.go.tz

Executive Director's Office
P. O Box 332,
**KIGOMA.**

---

**REF. NO: KDC/H/1/10**

May 13, 2025.

Medical Officer In-Charge,
Mwamgongo Health Center,
P.O. Box 555,
**KIGOMA.**

**RE: INTRODUCTION AND PERMISSION FOR DATA COLLECTION ON OUTCOME EVALUATION OF THE M-MAMA EMERGENCY TRANSPORT SYSTEM (EMTS) AND FACTORS ASSOCIATED WITH ITS UTILISATION AMONG LACTATING MOTHERS IN KIGOMA DISTRICT COUNCIL**

Please refer to the subject mentioned above.

2. Through this letter, I would like to introduce to you **Mr. Julius Masaba** from Mzumbe University, who is conducting a study titled: "*Outcome evaluation of the M-Mama emergency transport system (EmTS) and factors associated with its utilisation among lactating mothers in Kigoma District Council.*"

3. The purpose of this study is to examine the factors associated with the use of the M-Mama emergency transport system in improving the health of mothers and newborns. The researcher will collect data from women who delivered at the health facility and from those attending the MCH clinic at your facility, starting from the date of this letter until 30rd August 2025.

4. Kindly provide full cooperation to this researcher. Also, instruct the staff to offer all necessary assistance to facilitate the smooth completion of the study.

5. I wish you successful implementation.

..................

DISTRICT EXECUTIVE DIRECTOR
P.O.BOX 332
KIGOMA

Chiriku H. Chilumba
**EXECUTIVE DIRECTOR**
**KIGOMA DISTRICT COUNCIL**

**CC:** Deputy Vice Chancellor (Academic, Research and Consultancy)
Mzumbe University
P. O Box 1
Morogoro, Tanzania

**THE UNITED REPUBLIC OF TANZANIA**
**PRESIDENT'S OFFICE**
**REGIONAL GOVERNMENTS AND LOCAL GOVERNMENTS**
**KIGOMA DISTRICT COUNCIL**


Tovuti: www.kigomadc.go.tz

Executive Director's Office
P. O Box 332,
**KIGOMA.**

---

**REF. NO: KDC/H/1/10** May 13, 2025.

Medical Officer In-Charge,
Nkungwe Dispensary,
P.O. Box 555,
**KIGOMA.**

**RE: INTRODUCTION AND PERMISSION FOR DATA COLLECTION ON OUTCOME EVALUATION OF THE M-MAMA EMERGENCY TRANSPORT SYSTEM (EMTS) AND FACTORS ASSOCIATED WITH ITS UTILISATION AMONG LACTATING MOTHERS IN KIGOMA DISTRICT COUNCIL**

Please refer to the subject mentioned above.

2. Through this letter, I would like to introduce to you **Mr. Julius Masaba** from Mzumbe University, who is conducting a study titled: "*Outcome evaluation of the M-Mama emergency transport system (EmTS) and factors associated with its utilisation among lactating mothers in Kigoma District Council.*"

3. The purpose of this study is to examine the factors associated with the use of the M-Mama emergency transport system in improving the health of mothers and newborns. The researcher will collect data from women who delivered at the health facility and from those attending the MCH clinic at your facility, starting from the date of this letter until 30rd August 2025.

4. Kindly provide full cooperation to this researcher. Also, instruct the staff to offer all necessary assistance to facilitate the smooth completion of the study.

5. I wish you successful implementation.

………………
Chiriku H. Chilumba
**EXECUTIVE DIRECTOR**
**KIGOMA DISTRICT COUNCIL**

DISTRICT EXECUTIVE DIRECTOR
P.O.BOX 332
KIGOMA

**CC:** Deputy Vice Chancellor (Academic, Research and Consultancy)
Mzumbe University
P. O Box 1
Morogoro, Tanzania

**THE UNITED REPUBLIC OF TANZANIA**
**PRESIDENT'S OFFICE**
**REGIONAL GOVERNMENTS AND LOCAL GOVERNMENTS**
**KIGOMA DISTRICT COUNCIL**


Tovuti: www.kigomadc.go.tz

Executive Director's Office
P. O Box 332,
**KIGOMA.**

**REF. NO: KDC/H/1/10** May 13, 2025.

Medical Officer In-Charge,
Nyamhoza Dispensary,
P.O. Box 555,
**KIGOMA.**

**RE: INTRODUCTION AND PERMISSION FOR DATA COLLECTION ON OUTCOME EVALUATION OF THE M-MAMA EMERGENCY TRANSPORT SYSTEM (EMTS) AND FACTORS ASSOCIATED WITH ITS UTILISATION AMONG LACTATING MOTHERS IN KIGOMA DISTRICT COUNCIL**

Please refer to the subject mentioned above.

2. Through this letter, I would like to introduce to you **Mr. Julius Masaba** from Mzumbe University, who is conducting a study titled: "*Outcome evaluation of the M-Mama emergency transport system (EmTS) and factors associated with its utilisation among lactating mothers in Kigoma District Council.*"

3. The purpose of this study is to examine the factors associated with the use of the M-Mama emergency transport system in improving the health of mothers and newborns. The researcher will collect data from women who delivered at the health facility and from those attending the MCH clinic at your facility, starting from the date of this letter until 30rd August 2025.

4. Kindly provide full cooperation to this researcher. Also, instruct the staff to offer all necessary assistance to facilitate the smooth completion of the study.

5. I wish you successful implementation.

…………………
Chiriku H. Chilumba
**EXECUTIVE DIRECTOR**
**KIGOMA DISTRICT COUNCIL**

DISTRICT EXECUTIVE DIRECTOR
P.O.BOX 332
KIGOMA

**CC:** Deputy Vice Chancellor (Academic, Research and Consultancy)
Mzumbe University
P. O Box 1
Morogoro, Tanzania

**THE UNITED REPUBLIC OF TANZANIA**
**PRESIDENT'S OFFICE**
**REGIONAL GOVERNMENTS AND LOCAL GOVERNMENTS**
**KIGOMA DISTRICT COUNCIL**


Tovuti: www.kigomadc.go.tz

Executive Director's Office
P. O Box 332,
**KIGOMA.**

**REF. NO: KDC/H/1/10** May 13, 2025.

Medical Officer In-Charge,
Pamila Dispensary,
P.O. Box 555,
**KIGOMA.**

**RE: INTRODUCTION AND PERMISSION FOR DATA COLLECTION ON OUTCOME EVALUATION OF THE M-MAMA EMERGENCY TRANSPORT SYSTEM (EMTS) AND FACTORS ASSOCIATED WITH ITS UTILISATION AMONG LACTATING MOTHERS IN KIGOMA DISTRICT COUNCIL**

Please refer to the subject mentioned above.

2. Through this letter, I would like to introduce to you **Mr. Julius Masaba** from Mzumbe University, who is conducting a study titled: "*Outcome evaluation of the M-Mama emergency transport system (EmTS) and factors associated with its utilisation among lactating mothers in Kigoma District Council.*"

3. The purpose of this study is to examine the factors associated with the use of the M-Mama emergency transport system in improving the health of mothers and newborns. The researcher will collect data from women who delivered at the health facility and from those attending the MCH clinic at your facility, starting from the date of this letter until 30rd August 2025.

4. Kindly provide full cooperation to this researcher. Also, instruct the staff to offer all necessary assistance to facilitate the smooth completion of the study.

5. I wish you successful implementation.

..................
Chiriku H. Chilumba
**EXECUTIVE DIRECTOR**
**KIGOMA DISTRICT COUNCIL**

DISTRICT EXECUTIVE DIRECTOR
P.O.BOX 332
KIGOMA

**CC:** Deputy Vice Chancellor (Academic, Research and Consultancy)
Mzumbe University
P. O Box 1
Morogoro, Tanzania

THE UNITED REPUBLIC OF TANZANIA
PRESIDENT'S OFFICE
REGIONAL GOVERNMENTS AND LOCAL GOVERNMENTS
KIGOMA DISTRICT COUNCIL


Tovuti: www.kigomadc.go.tz

Executive Director's Office
P. O Box 332,
**KIGOMA.**

**REF. NO: KDC/H/1/10**

May 13, 2025.

Medical Officer In-Charge,
Chankele Dispensary,
P.O. Box 555,
**KIGOMA.**

**RE: INTRODUCTION AND PERMISSION FOR DATA COLLECTION ON OUTCOME EVALUATION OF THE M-MAMA EMERGENCY TRANSPORT SYSTEM (EMTS) AND FACTORS ASSOCIATED WITH ITS UTILISATION AMONG LACTATING MOTHERS IN KIGOMA DISTRICT COUNCIL**

Please refer to the subject mentioned above.

2. Through this letter, I would like to introduce to you **Mr. Julius Masaba** from Mzumbe University, who is conducting a study titled: "*Outcome evaluation of the M-Mama emergency transport system (EmTS) and factors associated with its utilisation among lactating mothers in Kigoma District Council.*"

3. The purpose of this study is to examine the factors associated with the use of the M-Mama emergency transport system in improving the health of mothers and newborns. The researcher will collect data from women who delivered at the health facility and from those attending the MCH clinic at your facility, starting from the date of this letter until 30rd August 2025.

4. Kindly provide full cooperation to this researcher. Also, instruct the staff to offer all necessary assistance to facilitate the smooth completion of the study.

5. I wish you successful implementation.

DISTRICT EXECUTIVE DIRECTOR
P.O.BOX 332
KIGOMA

..................
Chiriku H. Chilumba
**EXECUTIVE DIRECTOR**
**KIGOMA DISTRICT COUNCIL**

**CC:** Deputy Vice Chancellor (Academic, Research and Consultancy)
Mzumbe University
P. O Box 1
Morogoro, Tanzania

**THE UNITED REPUBLIC OF TANZANIA**
**PRESIDENT'S OFFICE**
**REGIONAL GOVERNMENTS AND LOCAL GOVERNMENTS**
**KIGOMA DISTRICT COUNCIL**


Tovuti: www.kigomadc.go.tz

Executive Director's Office
P. O Box 332,
**KIGOMA.**

**REF. NO: KDC/H/1/10** May 13, 2025.

Medical Officer In-Charge,
Kidahwe Dispensary,
P.O. Box 555,
**KIGOMA.**

**RE: INTRODUCTION AND PERMISSION FOR DATA COLLECTION ON OUTCOME EVALUATION OF THE M-MAMA EMERGENCY TRANSPORT SYSTEM (EMTS) AND FACTORS ASSOCIATED WITH ITS UTILISATION AMONG LACTATING MOTHERS IN KIGOMA DISTRICT COUNCIL**

Please refer to the subject mentioned above.

2. Through this letter, I would like to introduce to you **Mr. Julius Masaba** from Mzumbe University, who is conducting a study titled: "*Outcome evaluation of the M-Mama emergency transport system (EmTS) and factors associated with its utilisation among lactating mothers in Kigoma District Council.*"

3. The purpose of this study is to examine the factors associated with the use of the M-Mama emergency transport system in improving the health of mothers and newborns. The researcher will collect data from women who delivered at the health facility and from those attending the MCH clinic at your facility, starting from the date of this letter until 30rd August 2025.

4. Kindly provide full cooperation to this researcher. Also, instruct the staff to offer all necessary assistance to facilitate the smooth completion of the study.

5. I wish you successful implementation.

..................
Chiriku H. Chilumba
**EXECUTIVE DIRECTOR**
**KIGOMA DISTRICT COUNCIL**

DISTRICT EXECUTIVE DIRECTOR
P.O.BOX 332
KIGOMA

**CC:** Deputy Vice Chancellor (Academic, Research and Consultancy)
Mzumbe University
P. O Box 1
Morogoro, Tanzania
