## Supplementary material for "Outcome evaluation of the M-Mama emergency transport system (EmTS) and factors associated with its utilisation among lactating mothers in Kigoma District Council, Tanzania: A cross-sectional study": Questionnaire Table

### S2 Table. QUESTIONNAIRE USED FOR DATA COLLECTION IN KIGOMA RURAL COUNCIL, TANZANIA, 2025. (ENGLISH VERSION)

(√) Tick Answers

| QN | QUESTIONS | RESPONSE OPTIONS |
|---|---|---|
| **SECTION A: SOCIOECONOMIC, DEMOGRAPHIC CHARACTERISTICS AND FACTORS THAT INFLUENCE UTILISATION OF THE M-MAMA EMERGENCY TRANSPORT SYSTEM *(Objective 1)*** | | |
| **01** | Age (years) | ☐ 15-25<br>☐ 26-36<br>☐ 37-47 |
| **02** | Parity (Number of livebirth) | ☐ 1-2<br>☐ 3-4<br>☐ 5 or more |
| **03** | Marital Status | ☐ Single<br>☐ Married |
| **04** | Household Income Level | ☐ Low Income<br>☐ High Income |
| **05** | Education Level | ☐ None<br>☐ Primary<br>☐ Secondary and Above |
| **06** | Occupation | ☐ Farmer<br>☐ Housewife<br>☐ Tailor<br>☐ Employed<br>☐ No Occupation<br>☐ Other (specify): _______ |
| **07** | Place of residence | ☐ Town<br>☐ Village |
| **08** | Distance to nearest RCH facility | ☐ 0–3 Km<br>☐ 4–6 Km<br>☐ More than 6 Km |
| **SECTION B: PROJECT ENABLING FACTORS THAT INFLUENCE THE UTILISATION OF M-MAMA EMERGENCY TRANSPORT SYSTEM *(Objective 2)*** | | |
| **09** | Are you aware of the M-Mama Emergency Transport System? | ☐ Yes<br>☐ No |
| **10** | Have you ever seen advertisements of M-Mama EmTS? | ☐ Yes<br>☐ No<br>☐ I don't Know |
| **11** | Where have you seen advertisements of M-Mama EmTS? | ☐ Community<br>☐ RCH<br>☐ Not Sure<br>☐ Other (specify): _______ |
| **12** | Is the obstetric and neonatal emergency care call center number of M-Mama easily reachable when at village? | ☐ Yes<br>☐ No<br>☐ I don't Know |
| **13** | Is obstetric and neonatal emergency care readily available at primary health facility? | ☐ Yes<br>☐ No<br>☐ I don't Know |

| SECTION C: PROPORTION OF UTILISATION OF THE M-MAMA EMERGENCY TRANSPORT SYSTEM *(Objective 3)* | | |
|---|---|---|
| **14** | How did you utilise emergency transport service? | ☐ Paid (Not Utilise-M-Mama)<br>☐ Free (Utilise M-Mama) |
| **15** | What type of vehicle did you use during obstetric or newborn health emergency? | ☐ Private Vehicle<br>☐ M-Mama Ambulance<br>☐ M-Mama Community Tax<br>☐ Other (specify): _______ |
| **16** | How did you arrange emergency transport for referral? | ☐ I arranged it myself<br>☐ Relative<br>☐ TBA<br>☐ Healthcare Provider<br>☐ Other (specify): _______ |
| **17** | How many times have you used the emergency transport for referral? | ☐ Once<br>☐ Twice |
| **18** | Why did you utilise or not utilise the M-Mama emergency transport? | ☐ Free Service<br>☐ Emergency<br>☐ Circumstances<br>☐ Quick Response<br>☐ Not Aware of M-Mama<br>☐ Other (specify): _______ |
| **SECTION D: EXPERIENCE, PERCEPTION AND ATTITUDES TOWARD THE M-MAMA EMERGENCY TRANSPORT SYSTEM** ***(Objective 4)*** | | |
| **19** | I believe M-Mama EmTS is effective in handling maternal and newborn emergencies. | ☐ Agree<br>☐ Disagree<br>☐ Neutral |
| **20** | Negative experiences on emergency services cause women to not utilizes the M-Mama services | ☐ Agree<br>☐ Disagree<br>☐ Neutral |
| **21** | I am satisfied with the services provided by M-Mama EmTS | ☐ Agree<br>☐ Disagree<br>☐ Neutral |
| **22** | I trust the M-Mama EmTS transport providers (e.g., drivers, dispatchers) | ☐ Agree<br>☐ Disagree<br>☐ Neutral |
| **23** | The M-Mama transport provides quick transport during emergencies all the time | ☐ Agree<br>☐ Disagree<br>☐ Neutral |
| **24** | Would recommend the M-Mama Services to other Women | ☐ Agree<br>☐ Disagree<br>☐ Neutral |
| **25** | RCH Facility Name | ☐ Bitale Health Center<br>☐ Bubango Dispensary<br>☐ Chankabwimba Dispensary<br>☐ Kagongo Dispensary<br>☐ Kagunga Dispensary<br>☐ Kalalangabo Dispensary<br>☐ Kamara Dispensary |

| | | |
|---|---|---|
| | | ☐ Kaseke Dispensary<br>☐ Kidahwe Dispensary<br>☐ Kiganza Dispensary<br>☐ Kimbwela Dispensary<br>☐ Mahembe Dispensary<br>☐ Matendo Dispensary<br>☐ Matyazo Health Center<br>☐ Mgaraganza Dispensary<br>☐ Mkongoro Dispensary<br>☐ Mwamgongo Health Center<br>☐ Nkungwe Dispensary<br>☐ Nyamhoza Dispensary<br>☐ Pamila Dispensary<br>☐ Chankele Dispensary |
