## Supplementary material for "Outcome evaluation of the M-Mama emergency transport system (EmTS) and factors associated with its utilisation among lactating mothers in Kigoma District Council, Tanzania: A cross-sectional study": Stata Do-file for Data Analysis: S4 Do File.do

```
*JULIUS MASABA M-MAMA*
*closing opened log automatic when running the do file*
*capture log close
*importing data*
*log using "E:\M-MAMA\DATA SET\M-MAMA LOG.smcl"
*save "E:\M-MAMA\DATA SET\Mama Data.dta"
describe
list in 1/10
cd "D:\HD 1\M-MAMA\DATA SET"

*DATA CLEANING AND MANAGEMENT*

*Generating numeric variables/STATA Machinery readable format*

generate Age=.
replace Age=0 if agegrp=="15-25"
replace Age=1 if agegrp=="26-36"
label define Age 0 "15-25" 1 "26-36"
label values Age Age
label variable Age "Ages"
codebook Age
drop agegrp


generate Parity=.
replace Parity=0 if parity=="1-2"
replace Parity=1 if parity=="3-4"
label define Parity 0 "1-2" 1 "3-4"
label values Parity Parity
label variable Parity "Number of Children"
codebook Parity
drop parity


generate Marital=.
replace Marital=0 if marital=="Married"
replace Marital=1 if marital=="Single"
label define Marital 0 "Married" 1 "Single"
label values Marital Marital
label variable Marital "Marital Status"
codebook Marital
drop marital

generate Income=.
replace Income=0 if income=="Low Income"
replace Income=1 if income=="High Income"
label define Income 0 "Low Income" 1 "High Income"
label values Income Income
label variable Income "Income Level"
codebook Income
drop income


generate Education=.
replace Education=0 if education=="None"
replace Education=1 if education=="Primary"
replace Education=2 if education=="Secondary+"
label define Education 0 "None" 1 "Primary" 2 "Secondary+"
label values Education Education
label variable Education "Education Level"
codebook Education
drop education


generate Occupation=.
replace Occupation=0 if occupation=="Farmer"
replace Occupation=1 if occupation=="Housewife"
replace Occupation=2 if occupation=="Tailor"
replace Occupation=3 if occupation=="Employed"
label define Occupation 0 "Farmer" 1 "Housewife" 2 "Tailor" 3 "Employed"
label values Occupation Occupation
label variable Occupation "Occupations"
codebook Occupation
drop occupation


generate Residence=.
replace Residence=0 if resident=="Village"
replace Residence=1 if resident=="Town"
label define Residence 0 "Village" 1 "Town"
label values Residence Residence
label variable Residence "Resident"
codebook Residence
drop resident


generate Distance=.
replace Distance=0 if distance=="4-6 Km"
replace Distance=1 if distance=="0-3 Km"
label define Distance 0 "4-6 Km" 1 "0-3 Km"
label values Distance Distance
label variable Distance "Distances"
codebook Distance
drop distance

generate Awareness=.
replace Awareness=0 if aware_m_mama=="No"
replace Awareness=1 if aware_m_mama=="Yes"
label define Awareness 0 "No" 1 "Yes"
label values Awareness Awareness
label variable Awareness "M-Mama Awareness"
codebook Awareness
drop aware_m_mama


generate MamaAdvert=.
replace MamaAdvert=0 if seen_advert=="No"
replace MamaAdvert=1 if seen_advert=="Yes"
label define MamaAdvert 0 "No" 1 "Yes"
label values MamaAdvert MamaAdvert
label variable MamaAdvert "M-Mama Advert"
codebook MamaAdvert
drop seen_advert


generate MamaSource=.
replace MamaSource=0 if advert_source=="Not Sure"
replace MamaSource=1 if advert_source=="Community"
replace MamaSource=2 if advert_source=="RCH"
label define MamaSource 0 "Not Sure" 1 "Community" 2 "RCH"
label values MamaSource MamaSource
label variable MamaSource "M-Mama AdvSource"
codebook MamaSource
drop advert_source


generate MamaNumber=.
replace MamaNumber=0 if number_reachable=="I don't Know"
replace MamaNumber=1 if number_reachable=="Yes"
replace MamaNumber=2 if number_reachable=="No"
label define MamaNumber 0 "I don't Know" 1 "Yes" 2 "No"
label values MamaNumber MamaNumber
label variable MamaNumber "M-Mama Number Reachable"
codebook MamaNumber
drop number_reachable


generate ObstetricNeonatal=.
replace ObstetricNeonatal=0 if obsteneoermgcyavailable=="No"
replace ObstetricNeonatal=1 if obsteneoermgcyavailable=="Yes"
label define ObstetricNeonatal 0 "No" 1 "Yes"
label values ObstetricNeonatal ObstetricNeonatal
label variable ObstetricNeonatal "OBS and NEO Emergency Available"
codebook ObstetricNeonatal
drop obsteneoermgcyavailable


generate Utilization=.
replace Utilization=0 if utilization=="Not Utilise M-Mama"
replace Utilization=1 if utilization=="Utilise M-Mama"
label define Utilization 0 "Not Utilise M-Mama" 1 "Utilise M-Mama"
label values Utilization Utilization
label variable Utilization "M-Mama Utilization"
codebook Utilization
drop utilization


generate Transportused=.
replace Transportused=0 if vehicle=="Private Transport"
replace Transportused=1 if vehicle=="M-Mama Ambulance"
replace Transportused=2 if vehicle=="M-Mama Community Tax"
label define Transportused 0 "Private Transport" 1 "M-Mama Ambulance" 2 "M-Mama Community Tax"
label values Transportused Transportused
label variable Transportused "Transport Used"
codebook Transportused
drop vehicle


generate ArrangeTrans=.
replace ArrangeTrans=0 if arrange=="Healthcare Provider"
replace ArrangeTrans=1 if arrange=="Relative"
replace ArrangeTrans=2 if arrange=="TBA"
label define ArrangeTrans 0 "Healthcare Provider" 1 "Relative" 2 "TBA"
label values ArrangeTrans ArrangeTrans
label variable ArrangeTrans "Arrange Transport"
codebook ArrangeTrans
drop arrange


generate NumberEmTuse=.
replace NumberEmTuse=0 if times=="Once"
replace NumberEmTuse=1 if times=="Twice"
label define NumberEmTuse 0 "Once" 1 "Twice"
label values NumberEmTuse NumberEmTuse 
label variable NumberEmTuse "Number of EMT Used"
codebook NumberEmTuse
drop times


generate Reason=.
replace Reason=0 if reasons=="Emergency Circumstances"
replace Reason=1 if reasons=="Not Aware of M-Mama"
replace Reason=2 if reasons=="Free Service"
replace Reason=3 if reasons=="Quick Response"
label define Reason 0 "Emergency Circumstances" 1 "Not Aware of M-Mama" 2 "Free Service" 3 "Quick Response"
label values Reason Reason
label variable Reason "Reasons Use or Not Use M-Mama"
codebook Reason
drop reasons


generate MamaEffective=.
replace MamaEffective=0 if m_mama_effectveness=="Disagree"
replace MamaEffective=1 if m_mama_effectveness=="Neutral"
replace MamaEffective=2 if m_mama_effectveness=="Agree"
label define MamaEffective 0 "Disagree" 1 "Neutral" 2 "Agree"
label values MamaEffective MamaEffective
label variable MamaEffective "M-Mama Effectiveness"
codebook MamaEffective
drop m_mama_effectveness


generate Experiences=.
replace Experiences=0 if negative_experience=="Agree"
replace Experiences=1 if negative_experience=="Disagree"
replace Experiences=2 if negative_experience=="Neutral"
label define Experiences 0 "Agree" 1 "Disagree" 2 "Neutral"
label values Experiences Experiences
label variable Experiences "Negative Experience"
codebook Experiences
drop negative_experience


generate Satisfactions=.
replace Satisfactions=0 if m_mama_satisfaction=="Neutral"
replace Satisfactions=1 if m_mama_satisfaction=="Agree"
replace Satisfactions=2 if m_mama_satisfaction=="Disagree"
label define Satisfactions 0 "Neutral" 1 "Agree" 2 "Disagree"
label values Satisfactions Satisfactions
label variable Satisfactions "M-Mama Satisfied"
codebook Satisfactions
drop m_mama_satisfaction


generate Trusts=.
replace Trusts=0 if m_mama_trust=="Disagree"
replace Trusts=1 if m_mama_trust=="Agree"
replace Trusts=2 if m_mama_trust=="Neutral"
label define Trusts 0 "Disagree" 1 "Agree" 2 "Neutral"
label values Trusts Trusts
label variable Trusts "M-Mama Trust"
codebook Trusts
drop m_mama_trust


generate Quicks=.
replace Quicks=0 if m_mama_quick=="Disagree"
replace Quicks=1 if m_mama_quick=="Agree"
replace Quicks=2 if m_mama_quick=="Neutral"
label define Quicks 0 "Disagree" 1 "Agree" 2 "Neutral"
label values Quicks Quicks
label variable Quicks "M-Mama Quick"
codebook Quicks
drop m_mama_quick


generate Recommends=.
replace Recommends=0 if m_mama_recommend=="Disagree"
replace Recommends=1 if m_mama_recommend=="Agree"
replace Recommends=2 if m_mama_recommend=="Neutral"
label define Recommends 0 "Disagree" 1 "Agree" 2 "Neutral"
label values Recommends Recommends
label variable Recommends "M-Mama Recommend"
codebook Recommends
drop m_mama_recommend


generate Facility=.
replace Facility=0 if facility_name=="Bitale"
replace Facility=1 if facility_name=="Bubango"
replace Facility=2 if facility_name=="Chankambwimba"
replace Facility=3 if facility_name=="Kagongo"
replace Facility=4 if facility_name=="Kagunga"
replace Facility=5 if facility_name=="Kalalangabo"
replace Facility=6 if facility_name=="Kamara"
replace Facility=7 if facility_name=="Kaseke"
replace Facility=8 if facility_name=="Kidahwe"
replace Facility=9 if facility_name=="Kiganza"
replace Facility=10 if facility_name=="Kimbwela"
replace Facility=11 if facility_name=="Mahembe"
replace Facility=12 if facility_name=="Matendo"
replace Facility=13 if facility_name=="Matyazo"
replace Facility=14 if facility_name=="Mgaranganza"
replace Facility=15 if facility_name=="Mkongoro"
replace Facility=16 if facility_name=="Mwamgongo"
replace Facility=17 if facility_name=="Nkungwe"
replace Facility=18 if facility_name=="Nyamhoza"
replace Facility=19 if facility_name=="Pamila"
replace Facility=20 if facility_name=="Chankele"
label define Facility 0 "Bitale" 1 "Bubango" 2 "Chankambwimba" 3 "Kagongo" 4 "Kagunga" 5 "Kalalangabo" 6 "Kamara" 7 "Kaseke" 8 "Kidahwe" 9 "Kiganza" 10 "Kimbwela" 11 "Mahembe" 12 "Matendo" 13 "Matyazo" 14 "Mgaranganza" 15 "Mkongoro" 16 "Mwamgongo" 17 "Nkungwe" 18 "Nyamhoza" 19 "Pamila" 20 "Chankele"
label values Facility Facility
label variable Facility "Facility Name"
codebook Facility
drop facility_name


*ANALYSIS FOR RESULTS*

*1. DEMOGRAPHIC AND SOCIOECONOMIC CHARACTERISTICS OF RESPONDENTS*
*DISCRIPTIVE ANALYSIS*//
tab Age
tab Parity
tab Marital
tab Distance
tab Education
tab Income
tab Occupation
tab Residence
graph bar (count), over(Education)
graph pie, over(Marital)
histogram Age


*2. FACTORS INFLUENCING THE UTILIZATION OF M-MAMA EMTS*
*DISCRIPTIVE ANALYSIS*//
tab Utilization
tab Age
tab Parity
tab Marital
tab Distance
tab Education
tab Income
tab Occupation
tab Residence
tab Awareness
tab MamaAdvert
tab MamaSource
tab MamaNumber
tab ObstetricNeonatal
graph bar (count), over(Utilization)
graph pie, over(MamaSource)
histogram MamaNumber
graph bar (count), over(Education)
graph pie, over(Income)
histogram Occupation

*INFERENTIAL ANALYSIS*//
*Cross-tabulation*
tab Age Utilization, column
tab Parity Utilization, column
tab Occupation Utilization, column
tab Education Utilization, column
tab Income Utilization, column
tab Distance Utilization, column
tab Marital Utilization, column
tab Residence Utilization, column
tab Awareness Utilization, column
tab MamaAdvert Utilization, column
tab MamaSource Utilization, column
tab MamaNumber Utilization, column
tab ObstetricNeonatal Utilization, column
graph bar (count), over(Utilization)
graph pie, over(MamaSource)
histogram MamaNumber
graph bar (count), over(Awareness) over(Utilization)

*Chi2*
tab Age Utilization, column chi2
tab Parity Utilization, column chi2
tab Occupation Utilization, column chi2
tab Education Utilization, column chi2
tab Income Utilization, column chi2
tab Distance Utilization, column chi2
tab Marital Utilization, column chi2
tab Residence Utilization, column chi2
tab Awareness Utilization, chi2 row
tab MamaAdvert Utilization, chi2 row
tab MamaSource Utilization, chi2 row
tab MamaNumber Utilization, chi2 row
tab ObstetricNeonatal Utilization, chi2 row
graph bar (count), over(Utilization)
graph pie, over(MamaSource)
histogram MamaNumber
graph bar (count), over(Awareness) over(Utilization)
graph bar (count), over(ObstetricNeonatal) over(Utilization)

*Bivariable Logistic Regression* (COR)
logistic Utilization i.Age 
logistic Utilization i.Parity
logistic Utilization i.Occupation
logistic Utilization i.Education 
logistic Utilization i.Income
logistic Utilization i.Distance
logistic Utilization i.Marital 
logistic Utilization i.Residence
logistic Utilization i.Awareness
logistic Utilization i.MamaAdvert
logistic Utilization i.MamaSource 
logistic Utilization i.MamaNumber
logistic Utilization i.ObstetricNeonatal
estat gof

*MULTIVARIABLE LOGISTIC REGRESSION ANALYSIS*//
*Multvariable Logistic Regression* (AOR)
logistic Utilization i.Parity i.Education i.Income i.Distance i.Residence i.Awareness 
estat gof


*3. PROPORTION OF UTILIZATION OF THE M-MAMA EMTS AMONG LACTATING MOTHERS*
*DISCRIPTIVE ANALYSIS*//
tab Utilization
tab Transportused
tab ArrangeTrans
tab NumberEmTuse
tab Reason
tab Facility
graph bar (count), over(Transportused)
graph bar (count), over(Utilization)
graph pie, over(Transportused)
graph pie, over(Reason)

*Cross-tabulation*
tab Utilization
tab Transportused Utilization, column
tab ArrangeTrans Utilization, column
tab NumberEmTuse Utilization, column
tab Reason Utilization, column
tab Facility Utilization, column

*4. PERCEPTION OF LACTATING MOTHERS IN THE UTILIZATION OF THE M-MAMA EMTS*
*DISCRIPTIVE ANALYSIS*//
tab MamaEffective
tab Experiences 
tab Satisfactions
tab Trusts
tab Quicks
tab Recommends

*BINARY ANALYSIS*//
*Cross-tabulation*
tab MamaEffective
tab Experiences Utilization
tab Satisfactions Utilization
tab Trusts Utilization
tab Quicks Utilization
tab Recommends Utilization

*Chi2*
tab MamaEffective
tab Satisfactions Utilization, chi2 row
tab Experiences Utilization, chi2 row
tab Trusts Utilization, chi2 row
tab Quicks Utilization, chi2 row
tab Recommends Utilization, chi2 row


*FINAL REGRESSION MODEL*
*stepwise*
stepwise, pr(0.05): logistic Utilization i.Age i.Parity i.Occupation i.Education i.Income i.Distance i.Marital i.Residence i.Awareness i.MamaAdvert i.MamaSource i.MamaNumber i.ObstetricNeonatal, or
estat gof
```
